# Post-COVID Condition and Disparities in Daily Functional Activities: Findings from Virus Watch - a prospective community cohort study

**DOI:** 10.1101/2025.08.26.25334469

**Authors:** Wing Lam Erica Fong, Sarah Beale, Vincent Grigori Nguyen, Jana Kovar, Alexei Yavlinsky, Andrew C Hayward, Ibrahim Abubakar, Sander MJ van Kuijk, Robert W Aldridge

## Abstract

**Background:** Post-COVID Condition (PCC) is increasingly recognised to impair daily functioning, particularly work ability, cognitive function, and self-care. This analysis investigates the role of deprivation, migration status, and ethnicity in experiencing limitations in six functional activities: work/education, concentration, self-care, caring for others, performing necessary activities outside the house, and engaging in enjoyable activities.

**Methods:** We analysed data from Virus Watch, a prospective community cohort study in England, identifying 776 individuals (≥18 years) with PCC between February 2020 and March 2024. We used logistic regression to assess how deprivation, migration and ethnic minority status were associated with the odds of experiencing each functional limitation, adjusting for sociodemographic variables.

**Findings:** Individuals with PCC in IMD 1 (the most deprived quintile) had higher adjusted odds of limitations in work/education, concentration, self-care, and doing necessary activities outside the house than those in IMD 5 (the least deprived) (adjusted odds ratio (aOR) range: 2·20–2·85). Those in IMD 2 also experienced increased odds of limitations in work/education and concentration compared to those in IMD 5. We found no evidence of associations between migration status or minority ethnicity with functional limitations among PCC individuals.

**Interpretation:** Our findings indicate that socioeconomic deprivation, rather than migration status or ethnicity, is the primary driver of functional limitations within this cohort. Functional limitations may perpetuate cycles of deprivation and further exacerbate health inequalities. Equitable access to rehabilitation and support services, alongside workplace and educational adaptations, is needed to address the functional limitations of those affected by PCC.

**Funding:** Medical Research Council; Wellcome Trust; European Union

**Research in context:** *Evidence before this study:* We searched PubMed and Web of Science for English-language articles indexed from March 2020 up to May 2025 on the association between deprivation, migration status, and ethnicity, and functional limitations of post-COVID condition (PCC). The search string ‘(“post-COVID condition” OR “long COVID” OR “post-acute COVID-19” OR “post-COVID syndrome” OR “post-acute sequelae of SARS-CoV-2” OR PCC) AND (deprivation OR “Index of Multiple Deprivation” OR IMD OR “socioeconomic status” OR “social determinants” OR “social inequality” OR migration OR “migration status” OR migrant OR immigrant OR ethnicity OR “ethnic minority” OR race OR “racial group”) AND (“functional limitation*” OR “functional impairment*” OR “activity limitation*” OR “participation restriction*” OR “functional status” OR “work ability” OR “work attendance” OR “school attendance” OR “work participation” OR “school participation” OR concentration OR “cognitive impairment” OR “brain fog” OR “self-care” OR “activities of daily living” OR ADL OR “caring for others” OR caregiving OR “leisure activities” OR “enjoyable activities” OR “social participation” OR “necessary activities outside the house” OR “instrumental activities of daily living” OR IADL)’ was used. The search identified several US-based studies reporting greater functional limitations among Black and Hispanic populations, but few data exist outside the US, and none addressed deprivation and migration status. We therefore excluded the search terms related to social determinants in a follow-up search to focus on the broader literature on functional limitations. Most studies focused on hospitalised populations and often lacked detail on specific functional activities. One UK-based study reported greater functional impairment, measured using the Work and Social Adjustment Scale, among individuals in the most deprived quintile compared to those in less deprived quintiles in the UK. However, the impact of migration status remains largely unexplored.

*Added value of this study:* To our knowledge, this is the first study to examine how deprivation, migration status, and ethnic minority status influence specific functional limitations among adults with PCC in England. By disaggregating specific daily activities, we found that individuals living in more deprived areas experience greater odds of limitations in work/education attendance or participation, concentration, self-care, and doing necessary activities outside the house compared to those living in the least deprived areas. In contrast, no disparities were observed for migrants or ethnic minorities. Our findings provide granular insights into the role of social determinants, particularly deprivation, in influencing the functional impacts of PCC, allowing the development of more targeted interventions and policy responses to support those most affected.

*Implications of all the available evidence:* Alongside existing evidence, our findings highlight the need for targeted interventions to address the disproportionate impact of PCC on individuals in deprived communities. Policy responses should prioritise equitable access to rehabilitation and support services through adapted referral pathways and culturally appropriate outreach. Reforming statutory sick pay eligibility and ensuring the availability of flexible workplace or educational arrangements should also be ensured for those affected. Addressing these systemic barriers will prevent further widening of health inequalities and support a more equitable recovery from the long-term effects of COVID-19, while also informing future public health strategies for post-acute conditions.

## Introduction

Post-COVID condition (PCC), or long COVID, is defined as the presence of new or persistent symptoms lasting more than two months and occurring within three months of an acute SARS-CoV-2 infection, without an alternative diagnosis^1^. These persistent symptoms, most commonly fatigue, shortness of breath, and cognitive dysfunction, can impact an individual’s ability to carry out everyday activities, such as attending work or education, and caring for themselves and others.^2,3^ Increasing evidence shows that PCC impairs daily functioning, particularly with work ability, cognitive function, and self-care. Longitudinal cohort studies and systematic reviews suggest that a significant proportion of people with PCC experience a decline in functional abilities months after infection.^4–7^ One meta-analysis reported that, across multiple countries, 13-55% of individuals with PCC were unable to return to work up to 12 months after infection, with many also experiencing increased absenteeism and reduced productivity.^4^ Cognitive dysfunction and memory issues are frequently reported in people with PCC, impacting their ability to concentrate, process information, and perform complex tasks.^2^ Beyond occupational and cognitive limitations, PCC is also associated with impairments in activities of daily living (ADLs), including both basic self-care tasks and instrumental ADLs such as managing household tasks, preparing meals and shopping, which can in turn reduce independence and quality of life.^8,9^

Although PCC imposes significant functional limitations on affected individuals across hospitalised and general populations, there is limited research on how socioeconomic deprivation, ethnicity, and migration status influence the functional outcomes of PCC. Emerging evidence suggests that individuals who are socioeconomically deprived may be at increased risk of experiencing functional limitations as a result of PCC. For example, a UK-based study found that individuals in the most deprived quintile had higher levels of functional impairment, as measured by the Work and Social Adjustment Scale, compared with those in less deprived quintiles.^10^ Similarly, several US-based studies observed greater cognitive impairments, including difficulty concentrating, among Black and Hispanic individuals hospitalised for COVID-19, but few data exist outside the US.^11–13^ Overall, the evidence for functional limitations among these populations is limited, but the risk of functional limitations, particularly reduced work/educational and self-care abilities, from PCC may reinforce and exacerbate existing health inequalities. Understanding these disparities is essential to inform equitable healthcare access, guide targeted rehabilitation and support services, and shape policy interventions to reduce the long-term impact of COVID-19 in these communities.

Thus, this study used data from Virus Watch, a community cohort study, to investigate how deprivation, migration status, and ethnicity are associated with experiencing six specific functional limitations in daily life: work/education attendance or participation, concentration, self-care, caring for others, performing necessary activities outside the house, and engaging in enjoyable activities.

## Methods

### Ethics Approval and Consent

Virus Watch was approved by the Hampstead NHS Health Research Authority Ethics Committee: 20/HRA/2320, and conformed to the ethical standards set out in the Declaration of Helsinki. Participants provided informed consent for all aspects of the study.

### Study design and participants

Virus Watch was a large prospective community cohort study conducted from June 2020 to March 2025 to examine the transmission and impact of COVID-19 in England and Wales, with 58,497 participants enrolled by March 2022. Participants self-selected into the study, with eligibility limited to households with internet access and a lead household member proficient in reading English, although consent forms were available in multiple languages. Participants completed weekly online surveys on COVID-19 symptoms, testing, and vaccination, along with occasional in-depth questionnaires on COVID-19-related topics like behavioural practices, healthcare access and long-term symptoms. Furthermore, the Virus Watch cohort was linked to Hospital Episode Statistics (HES) Admitted Patient Care, which contains details of all admissions at NHS hospitals in England and admissions to private or charitable hospitals paid for by the NHS.^14^ The Virus Watch dataset was also linked to the Second Generation Surveillance System (SGSS), the UK National Immunisation Management Service (NIMS) and mortality data from the Office for National Statistics (ONS). The full study design and methodology are described elsewhere.^15,16^

Participants in this analysis were a subset of the Virus Watch study cohort. They were included if they 1) were aged 18 years or above and registered with an English postcode, 2) were successfully linked to either HES, NIMS, SGSS or ONS mortality data, 3) were not hospitalised for/with COVID-19; and 4) met the World Health Organisation (WHO) consensus definition of PCC. Hospitalised individuals were excluded to prevent misclassifying cases of post-intensive-care or post-sepsis syndromes as PCC, and those aged below 18 years or residing in Wales were excluded due to the lack of linked hospitalisation data for these groups.

Following the WHO definition, PCC was defined as individuals with a confirmed SARS-CoV-2 infection within three months of symptom onset with at least one symptom lasting two months, unexplained by another diagnosis (ref). PCC cases were identified through six in-depth surveys on long-term symptoms administered across four years (February 2021, May 2021, March 2022, March 2023, October 2023, April 2024). In each survey, participants reported any symptoms lasting at least four weeks, the onset dates of their three most severe symptoms, and whether they were ongoing or resolved. Symptom duration was calculated from symptom onset to survey completion (for ongoing symptoms) or reported resolution date. Although data were collected from February 2020 to April 2024, only cases reported up to 31 March 2024 were included, as hospitalisation data were limited to this period.

### Exposures

We examined three exposures: socioeconomic deprivation, migration status, and minority ethnicity status.

Socioeconomic deprivation was measured using quintiles of the Index of Multiple Deprivation. IMD is calculated for small local areas in England and typically covers seven dimensions of deprivation: crime, employment, education, income, health, living environment, and barriers to housing and services. These areas are ranked from most to least deprived relative to others and categorised into five quintiles. The 1st quintile represents the most deprived areas, and the 5th quintile represents the least deprived. The IMD classification for each participant was determined based on their self-reported residential postcode in the baseline survey, which was linked to the May 2020 Office for National Statistics (ONS) Postcode Lookup dataset.^17^ For this analysis, the 5th quintile was the reference category.

Migration status was determined by self-reported country of birth, with individuals born outside the UK classified as migrants. The UK-born group served as the reference category.

Ethnic minority status was determined using self-reported ethnicity. Participants identifying as white Irish, white Other, mixed, South Asian, other Asian, Black, or other were categorised as ethnic minorities.^18^ The white British group was used as the reference category.

### Outcomes

Within our occasional surveys on persistent symptoms, participants who reported any symptoms lasting >4 weeks were asked for self-perceived limitations in their ability to: 1) go to/participate in work or education; 2) concentrate on things (e.g. reading, watching TV); 3) take care of themselves (e.g. wash, dress, and feed yourself); 4) take care of others in the household; 5) do necessary daily activities outside the household; and 6) do activities that you enjoy (e.g. hobbies). Participants were identified to have a specific functional limitation if they answered ‘Yes, a lot’ or ‘Yes, a little’. Those reporting ‘Not at all’ indicated they did not experience the specific functional limitation, while those reporting ‘Not applicable’ were excluded from the analysis for that functional limitation.

### Covariates

The covariates considered were age group (0-24, 25-44, 45-64, 65+ years), sex assigned at birth (male and female), and pre-infection health. Pre-infection health was assessed based on self-reported health conditions at baseline, with participants classified as ‘not clinically vulnerable’, ‘clinically vulnerable’, or ‘extremely clinically vulnerable’ according to the UK NHS/government criteria for clinical vulnerability.^19^

### Statistical Analysis

Baseline demographic and clinical characteristics were summarised using descriptive statistics.

We used logistic regression to investigate how deprivation, migration status, and ethnic minority status influenced the odds of experiencing each of the 6 functional limitations as a result of PCC. Results are expressed as odds ratios with 95% confidence intervals. Separate models were fitted for each exposure and functional limitation outcome. For models where migration status or ethnic minority status was the exposure, we conducted complete case analyses. However, for models where IMD was the exposure, participants with missing migration status were kept by including them in a separate “Missing” category, due to the high proportion of missingness (24%) for this variable.

We performed three sensitivity analyses. First, we conducted complete case analyses for models with IMD as the exposure to compare results with those from the main analysis, which included those with missing migration status. Second, we recalculated the IMD quintile measure, which excludes the health domain when calculating the ranking score, using the methodology described by Adams and White.^20^ We used this as the exposure to assess whether the inclusion of the health domain introduced endogeneity bias. Lastly, for analyses with migration status as the exposure, we included participants with missing country of birth in a separate ‘Missing’ category.

Potential confounders for each exposure were identified using directed acyclic graphs (Supplementary figures 1-6). Multicollinearity was assessed (see ‘Multicollinearity’ in Supplemental Materials). For deprivation, the minimal adjustment set included age, sex, ethnic minority status, and migration status. For migration status, the set consisted of age, sex, and ethnic minority status. For ethnic minority status, no confounders were identified, but we adjusted for age and sex based on *a priori* considerations. In our secondary analysis, we additionally adjusted for pre-infection health in models with deprivation and migration status as exposures, reflecting alternative assumptions about the directionality between pre-infection health and the exposures, i.e. reversing the direction of the relationship between pre-infection health and deprivation/migration status. Results from these models are presented in the supplementary materials.

We followed the STROBE guidelines for reporting the study (Supplementary Materials).

## Results

We identified 906 participants who met the WHO criteria for PCC. Thirty (3%) were excluded due to hospitalisation for or with COVID-19. 776 completed the functional limitations section of the survey, of which 686 (86%) reported experiencing at least one functional limitation related to PCC (Figure 1).

**Figure 1.**
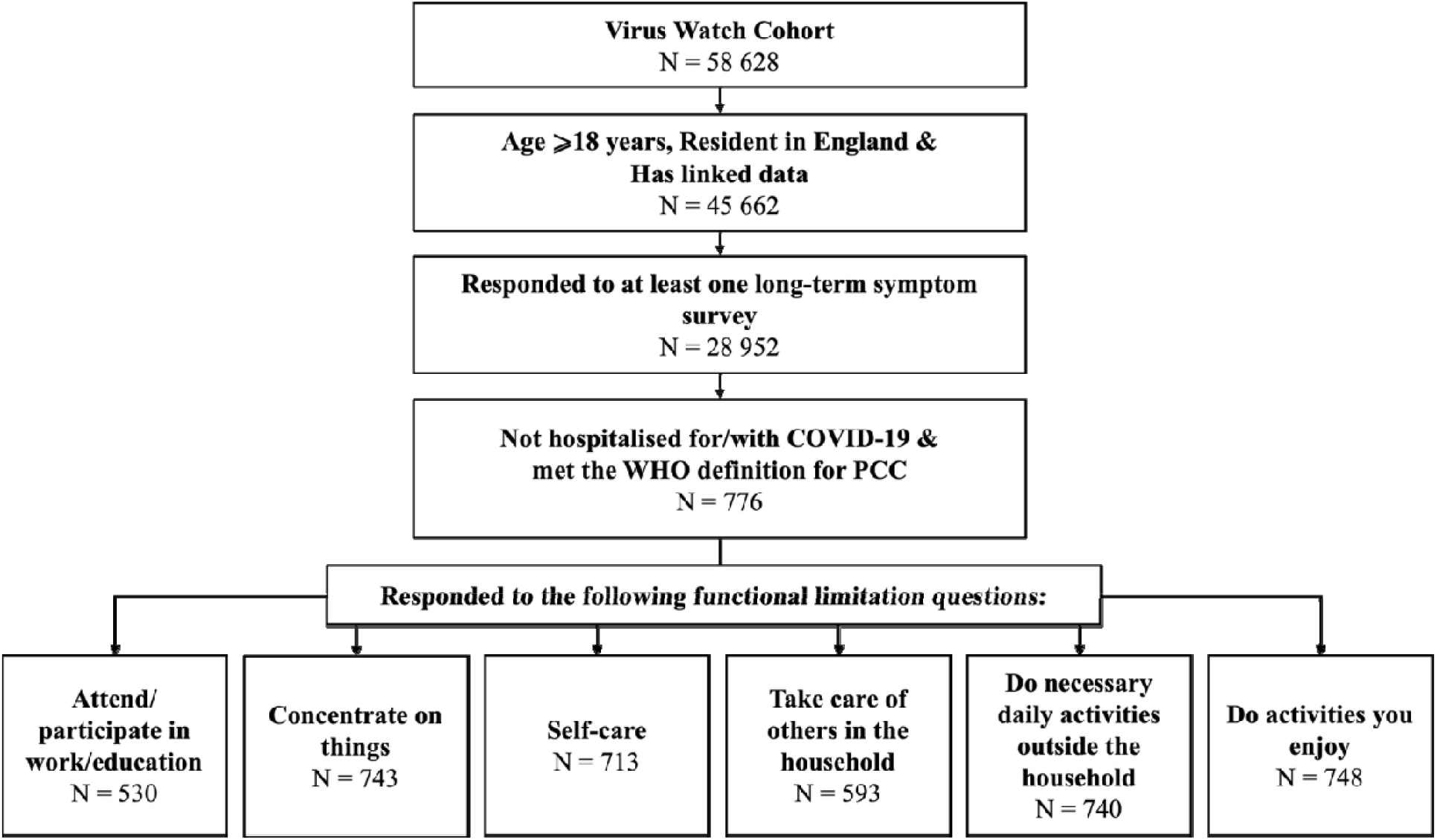
Cohort flow diagram.

Table 1 reports the sociodemographic and clinical characteristics of the analysis cohort. Overall, the majority within the cohort were 45-64 years of age (49%), female (71%), White British (88%), UK-born (69%), living in less deprived neighbourhoods (25%), and not clinically vulnerable (49%). Cohort characteristics by outcome and PCC status are presented in Supplementary Table 1. Participants answered the survey on functional limitations a median of 6.0 months (interquartile range = 6.7) after developing their first PCC symptom. High proportions of participants reported limitations in attending or participating in work or education (61%), concentrating (67%), and engaging in enjoyable activities (73%) (Table 2).

**Table 1.**
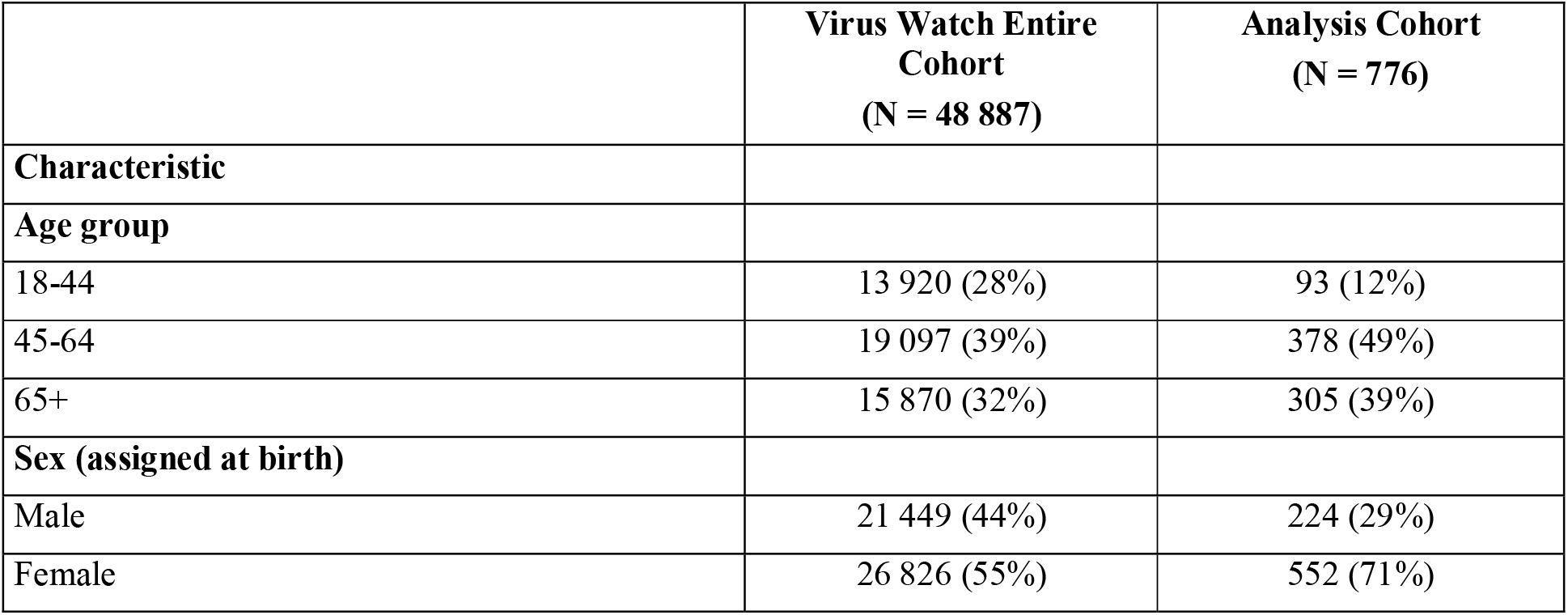
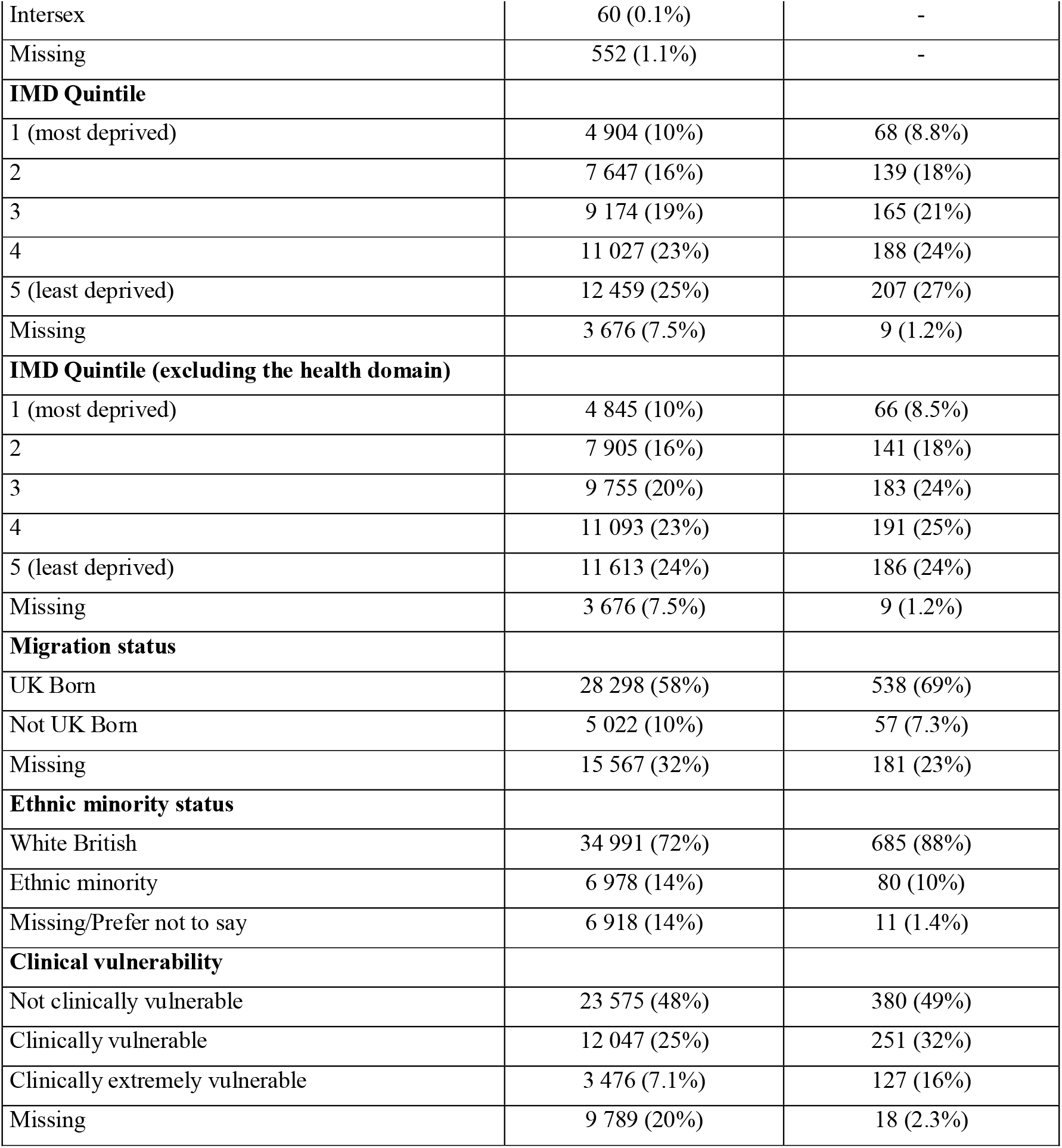
Sociodemographic and clinical characteristics of the analysis cohort compared to the entire Virus Watch study cohort.

**Table 2.** Number and proportion of participants who reported the presence or absence of limitations in each daily functional activity.

|  | Attend/<br>participate in<br>work/<br>education | Concentrate<br>on things | Self-care | Take care of<br>others in the<br>household | Do necessary<br>daily<br>activities<br>outside the<br>household | Do activities<br>you enjoy |
| --- | --- | --- | --- | --- | --- | --- |
|  | N = 530 | N = 743 | N = 713 | N = 593 | N = 740 | N = 748 |
| Yes | 325 (61%) | 495 (67%) | 186 (26%) | 198 (33%) | 448 (61%) | 545 (73%) |
| No | 205 (39%) | 248 (33%) | 527 (74%) | 395 (67%) | 292 (39%) | 203 (27%) |

### By IMD Quintile

Compared with participants in the least deprived (IMD 5) group, those in the most deprived IMD quintiles (1 and 2) had higher odds of experiencing limitations, after adjusting for age, sex, ethnic minority and migration status. Specifically, individuals in the most deprived quintile (IMD 1) had 2·46 [95% Confidence Interval (CI): 1·21–5·24] times greater odds of limitations in attending/participating in work/education and 2.85 [1·46–5·93] times higher odds of difficulties concentrating (Figure 2 & Supplementary tables 5-6). Participants in IMD 2 also had increased odds in these domains (work/education: 1.88 [1·09–3·29]; concentration: 1.85 [1·15–3·03]) compared to those in IMD 5.

**Figure 2.**
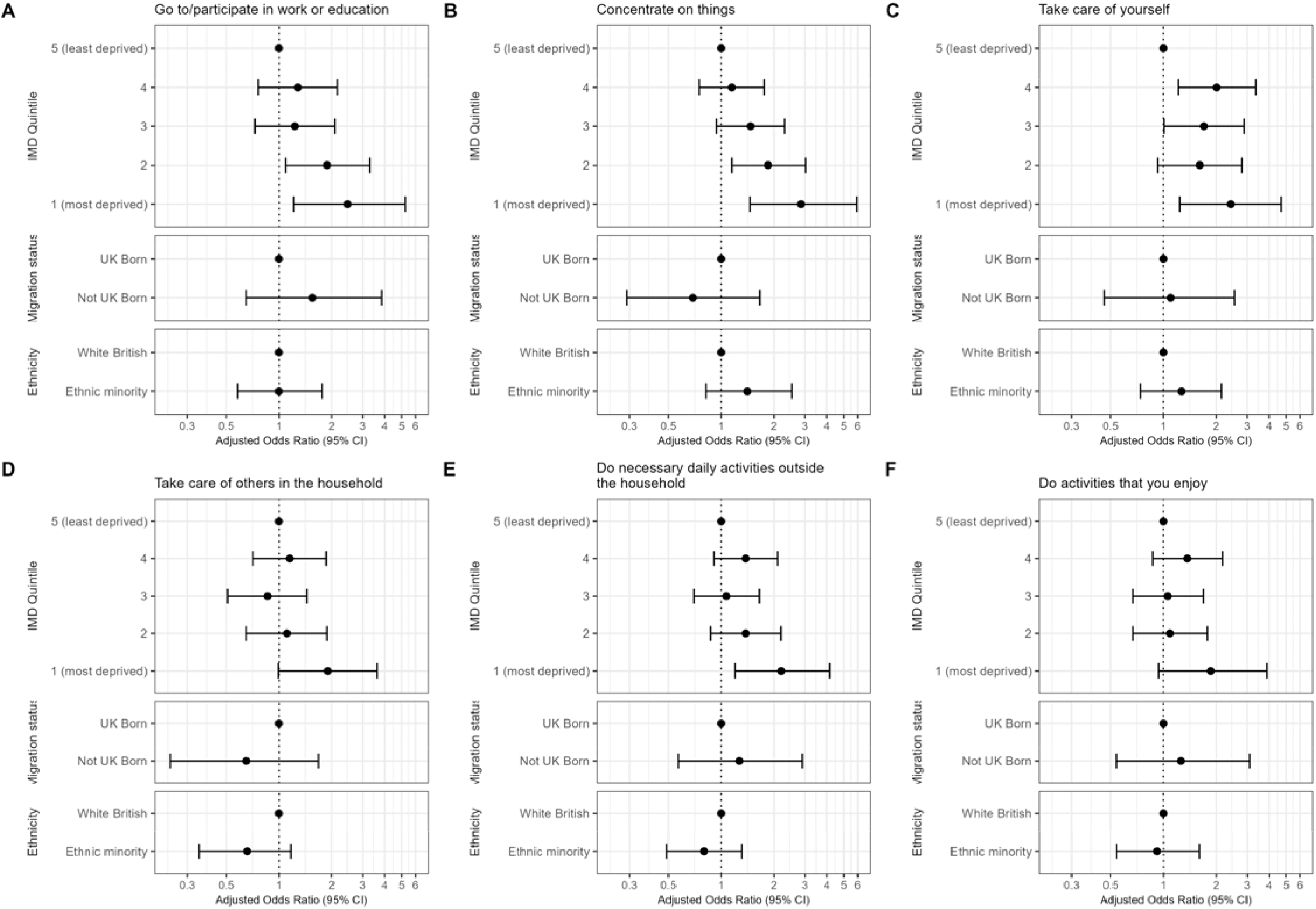
Forest plots of adjusted odds ratios of each exposure on limitations in A) attending or participating in work/education; B) concentrating; C) taking care of yourself; D) taking care of others in the household; E) doing necessary daily activities outside the household; and F) doing enjoyable activities.

Participants in the most deprived areas also had 2·42 (1·24–4·69) times greater odds of self-care limitations and 2·20 (1·20–4·15) times higher odds of difficulties performing necessary activities outside the house compared to the least deprived group (Figure 2 & Supplementary tables 7 & 9). Additionally, there was some evidence of increased odds of limitations in caring for others (1·90 [0·99–3·62]) and in engaging in enjoyable activities (1·86 [0·94–3·89]) among participants in the most deprived areas compared to the least deprived areas (Figure 2 & Supplementary tables 8 & 10).

The associations between area-level deprivation (IMD) and limitations in work/education attendance or participation, and concentration persisted after further adjusting for pre-infection health status. However, the effect of area-level deprivation on self-care activities, caring responsibilities, and engagement in enjoyable activities was attenuated (Supplementary Tables 5-10).

In the sensitivity analysis using IMD quintile excluding the health domain, associations remained consistent with those of the main analysis (Supplementary Tables 23-28). In the complete case sensitivity analysis, most associations were attenuated, with wider confidence intervals that included 1, except for the association with limitations in concentration, which remained. However, the direction of associations remained consistent across all outcomes (Supplementary Tables 29-34).

### By Migration Status

No association was observed between migration status and experiencing limitations in any functional activities after accounting for age, sex, and minority ethnic status (Figure 1; Supplementary Tables 11-16). This finding persisted following further adjustment for pre-infection health status and also in the sensitivity analyses, which included participants with unknown country of birth under “Missing” (Supplementary Tables 35-40).

### By Ethnic Minority Status

Similarly, after adjusting for age and sex, our findings indicated a lack of evidence for a difference in the odds of experiencing functional limitations between White British and ethnic minority participants.

## Discussion

Using data from a community cohort in England, we observed that people living in more deprived areas had increased odds of experiencing functional limitations related to PCC, specifically attending or participating in work/education, concentrating, self-care, and doing daily activities outside the household, compared to those in the least deprived areas. In contrast, the likelihood of experiencing functional limitations was similar between migrants and non-migrants, as well as between ethnic minorities and White British participants. These findings indicate that socioeconomic deprivation, rather than migration status or ethnicity, is the primary driver of functional limitations within this cohort.

Our findings are consistent with previous literature, indicating a greater likelihood of experiencing limitations in attending work or education due to PCC among individuals living in more deprived areas than those in the least deprived areas. Similarly, our results regarding migration status and experiencing functional limitations broadly align with those from a South London community study, which reported increased odds of functional limitations due to poor mental health among migrants compared to non-migrants, but no difference in limitations related to physical health problems.^21^ However, it is important to note that their study explored general functional limitations related to mental and physical health, rather than those specifically associated with PCC. While not directly comparable, both studies suggest that migration status alone may not be strongly associated with functional limitations resulting from physical health conditions. Furthermore, our results provide limited support for an association between ethnic minority status and cognitive limitations, a relationship previously reported by multiple US-based studies.^22–24^ Our results suggest a possible increased likelihood of concentration difficulties among ethnic minority participants with PCC. However, further research in a larger cohort is needed, as our analysis was limited by power.

Deprived communities may have a higher likelihood of experiencing functional limitations due to a range of systemic barriers. Reduced access to rehabilitation services is a key structural barrier, with evidence suggesting that people from socio-economically deprived areas were less likely to be referred to PCC-related services.^25^ Barriers, such as lack of awareness, fear of stigma, and distrust of healthcare providers, may further reduce service uptake in these populations. Furthermore, people with long-term conditions who are unable to access rehabilitation services are less likely to attend or participate in work/education.^26^ While some employers and educational institutions are beginning to offer more flexible arrangements in response to PCC, many individuals, particularly those in low-paid, insecure jobs, do not have access to flexible hours, extended rest breaks, remote work or learning options, or phased returns. Additionally, statutory sick pay in the UK is only available to employees earning above the income threshold, meaning that inadequate sick pay may compel individuals with PCC to return to work despite feeling unwell.^27^

These structural barriers make it difficult for affected individuals to manage persistent symptoms and remain in or return to work or education, contributing to prolonged absences and reduced participation in work or educational settings. These limitations can have far-reaching consequences, including financial insecurity, reduced quality of life, and poorer mental health. Financial strain and psychological distress may further impact functional status, limiting the ability to perform other daily activities like self-care, caregiving, and doing enjoyable activities.^28^ Ultimately, disruptions in work or education can perpetuate cycles of deprivation and exacerbate existing health inequalities, underscoring the need to address these systemic barriers to support recovery and minimise the broader social and economic impacts of PCC.

Policy responses should prioritise equitable access to and targeted rehabilitation services, such as adapted referral pathways, extended hours, and community outreach, that will address the full spectrum of PCC-related limitations.^29^ Targeted and culturally appropriate public health campaigns are also essential to raise awareness and reduce stigma around PCC and promote the availability of rehabilitation services. Additionally, workplace and educational institutions should be required to provide flexible arrangements, including remote options and occupational health support, to facilitate continued participation from those affected. Educating employers on PCC can support the implementation of better workplace adaptations to improve job and education retention. Statutory sick pay eligibility should also be reformed to support all workers, including those in low-paid, precarious jobs, ensuring adequate financial support during periods of illness and rehabilitation. Implementing these measures will not only help mitigate the long-term social and economic consequences of PCC, it will also help prevent widening health inequalities and support a more equitable recovery for both current and future public health emergencies.^30^

### Strengths and Limitations

Existing studies on functional impairments from PCC have predominantly focused on hospitalised populations, with limited research in community-based cohorts. To our knowledge, this is the first study to investigate the relationships between deprivation, migration status, ethnic minority status, and limitations in specific daily functioning activities resulting from PCC among community-based COVID-19 cases. Furthermore, using a detailed survey on functional limitations allowed us to disaggregate impacts across a range of daily activities, providing a more nuanced understanding of how PCC affects different population groups. These findings provided insights into the role of social determinants in shaping the functional impacts of PCC and can inform targeted interventions and policy responses.

However, this study had several limitations. Virus Watch participants were not fully representative of the English population, with an underrepresentation of individuals from more deprived areas, migrants, and ethnic minorities. This was likely due to language barriers, limited internet access, and economic circumstances, which may affect the generalisability of our findings and reduce statistical power. The inability to stratify ethnicity and migration status into more granular groups means that heterogeneity within these populations could not be examined, and further research with larger, diverse samples is needed to address this. The sample size was also insufficient to examine associations with self-perceived severity of functional limitations or the intersectionality between deprivation, migration status, and ethnicity. Some individuals with PCC may not have been identified due to limited community testing, recall bias, and undetected asymptomatic infections. Further recall bias may be introduced through the self-reporting of functional limitations. In addition, the use of IMD, an area-level measure of deprivation, may not accurately reflect individual socioeconomic position; future studies should use individual-level indicators, such as occupation and income. Lastly, since we did not use the PCFS scale to assess functional limitations, our results are not directly comparable with studies that did.

## Conclusion

The current study found evidence of elevated odds of experiencing functional limitations, particularly attending or participating in work/education, concentrating, self-care, and doing necessary activities outside the household, among people living in more deprived areas compared to those in the least deprived areas. Our findings underscore the need for equitable access to rehabilitation and support services and policies to ensure that workplaces and educational institutions adapt to the needs of those affected by PCC. Further investigation into more granular ethnicity and migration status groups, alongside the intersectionality of multiple social dimensions on long-term functional impacts on PCC is recommended to better inform targeted public health efforts, resource allocation, and policy responses for those most affected.

## Data Availability

We aim to share aggregate data from this project on our website and via a "Findings so far" section on our website - https://ucl-virus-watch.net/. We will also be sharing individual record-level data on a research data sharing service such as the Office for National Statistics Secure Research Service. In sharing the data, we will work within the principles set out in the UKRI Guidance on best practice in the management of research data. Access to use of the data whilst research is being conducted will be managed by the Chief Investigators (AH, RA and IA) in accordance with the principles set out in the UKRI guidance on best practice in the management of research data. We will put analysis code on publicly available repositories to enable their reuse.

## Acknowledgements

We would like to thank all the members of the END-VoC Long COVID advisory group for their input on the conceptualisation of this study.

## Contributors

Conceptualisation (WLEF, SB, SvK, RA), Data curation (WLEF, SB, VN, AY), Formal Analysis (WLEF), Funding acquisition (RA, AH, IA), Methodology (WLEF, SB, SvK, RA), Project administration (JK), Software (VN), Supervision (SB, SvK RA), Validation (VN, SB), Visualisation (WLEF), Writing – original draft (WLEF), Writing – reviewing and editing (all)

