## Supplementary Materials for "Post-COVID Condition and Disparities in Daily Functional Activities: Findings from Virus Watch - a prospective community cohort study"

### **Contents**

**STROBE checklist**………………………………………..……………………………………………………………………………………………………………….. 2

**Directed Acyclic Graphs**

**Sociodemographic and Clinical Characteristics**

**Multicollinearity**

**Main Analyses - Unadjusted and Adjusted Odds Ratios Exposure: IMD Quintile as Exposure**

**Main Analyses - Unadjusted and Adjusted Odds Ratios Exposure: Migration Status as Exposure**

**Main Analyses - Unadjusted and Adjusted Odds Ratios Exposure: Ethnic Minority Status as Exposure**

**Sensitivity Analyses 1 - Unadjusted and Adjusted Odds Ratios Exposure: IMD Quintile as Exposure**

**Sensitivity Analyses 2 - Unadjusted and Adjusted Odds Ratios Exposure: IMD Quintile as Exposure**

**Sensitivity Analyses 3 - Unadjusted and Adjusted Odds Ratios Exposure: Migration Status (including the 'Missing' category) as Exposure**

**STROBE Statement**

|  | Item No | Recommendation | Page No |
| --- | --- | --- | --- |
| **Title and abstract** | 1 | (*a*) Indicate the study’s design with a commonly used term in the title or the abstract | 2 |
|  |  | (*b*) Provide in the abstract an informative and balanced summary of what was done and what was found |  |
| Introduction | | | |
| Background/rationale | 2 | Explain the scientific background and rationale for the investigation being reported | 4 |
| Objectives | 3 | State specific objectives, including any prespecified hypotheses | 4 |
| Methods | | | |
| Study design | 4 | Present key elements of study design early in the paper | 4, 5 |
| Setting | 5 | Describe the setting, locations, and relevant dates, including periods of recruitment, exposure, follow-up, and data collection | 4, 5 |
| Participants | 6 | (*a*) Give the eligibility criteria, and the sources and methods of selection of participants. Describe methods of follow-up | 5 |
|  |  | (*b*) For matched studies, give matching criteria and number of exposed and unexposed |  |
| Variables | 7 | Clearly define all outcomes, exposures, predictors, potential confounders, and effect modifiers. Give diagnostic criteria, if applicable | 5, 6 |
| Data sources/ measurement | 8* | For each variable of interest, give sources of data and details of methods of assessment (measurement). Describe comparability of assessment methods if there is more than one group | 5, 6 |
| Bias | 9 | Describe any efforts to address potential sources of bias | 5 |
| Study size | 10 | Explain how the study size was arrived at | 5 |
| Quantitative variables | 11 | Explain how quantitative variables were handled in the analyses. If applicable, describe which groupings were chosen and why | 5, 6 |
| Statistical methods | 12 | (*a*) Describe all statistical methods, including those used to control for confounding | 6 |
|  |  | (*b*) Describe any methods used to examine subgroups and interactions |  |
|  |  | (*c*) Explain how missing data were addressed |  |
|  |  | (*d*) If applicable, explain how loss to follow-up was addressed |  |
|  |  | (*e*) Describe any sensitivity analyses |  |
| Results | | |  |
| Participants | 13* | (a) Report numbers of individuals at each stage of study—eg numbers potentially eligible, examined for eligibility, confirmed eligible, included in the study, completing follow-up, and analysed | 7, Fig 1 |
|  |  | (b) Give reasons for non-participation at each stage |  |
|  |  | (c) Consider use of a flow diagram |  |
| Descriptive data | 14* | (a) Give characteristics of study participants (eg demographic, clinical, social) and information on exposures and potential confounders | 7, 8 |
|  |  | (b) Indicate number of participants with missing data for each variable of interest |  |
|  |  | (c) Summarise follow-up time (eg, average and total amount) |  |
| Outcome data | 15* | Report numbers of outcome events or summary measures over time | 7, Fig 1 |

| Main results | 16 | (*a*) Give unadjusted estimates and, if applicable, confounder-adjusted estimates and their precision (eg, 95% confidence interval). Make clear which confounders were adjusted for and why they were included | 8, 9, 10 |
| --- | --- | --- | --- |
|  |  | (*b*) Report category boundaries when continuous variables were categorized |  |
|  |  | (*c*) If relevant, consider translating estimates of relative risk into absolute risk for a meaningful time period |  |
| Other analyses | 17 | Report other analyses done—eg analyses of subgroups and interactions, and sensitivity analyses | 9 |
| Discussion | | | |
| Key results | 18 | Summarise key results with reference to study objectives | 11 |
| Limitations | 19 | Discuss limitations of the study, taking into account sources of potential bias or imprecision. Discuss both direction and magnitude of any potential bias | 12 |
| Interpretation | 20 | Give a cautious overall interpretation of results considering objectives, limitations, multiplicity of analyses, results from similar studies, and other relevant evidence | 11, 12 |
| Generalisability | 21 | Discuss the generalisability (external validity) of the study results | 12 |
| Other information | | | |
| Funding | 22 | Give the source of funding and the role of the funders for the present study and, if applicable, for the original study on which the present article is based | 12 |

*Give information separately for exposed and unexposed groups.

**Note:** An Explanation and Elaboration article discusses each checklist item and gives methodological background and published examples of transparent reporting. The STROBE checklist is best used in conjunction with this article (freely available on the Web sites of PLoS Medicine at http://www.plosmedicine.org/, Annals of Internal Medicine at http://www.annals.org/, and Epidemiology at http://www.epidem.com/). Information on the STROBE Initiative is available at http://www.strobe-statement.org.

### **Directed Acyclic Graph**

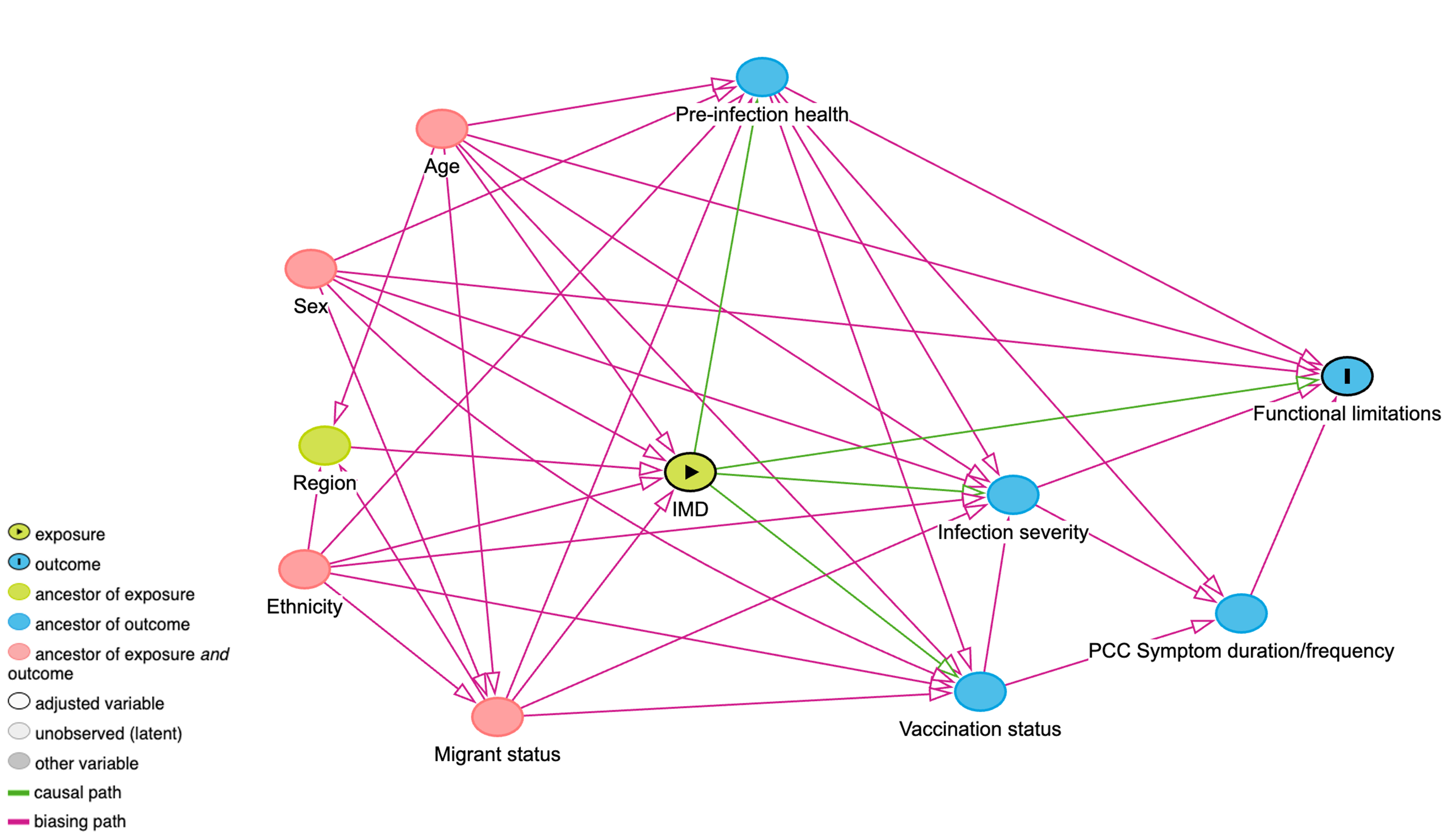

**Figure 1 Directed Acyclic Graph for the impact of deprivation (IMD Quintile) on experiencing functional limitations among people with PCC. This adjustment set holds the assumptions of deprivation influencing pre-infection health.**

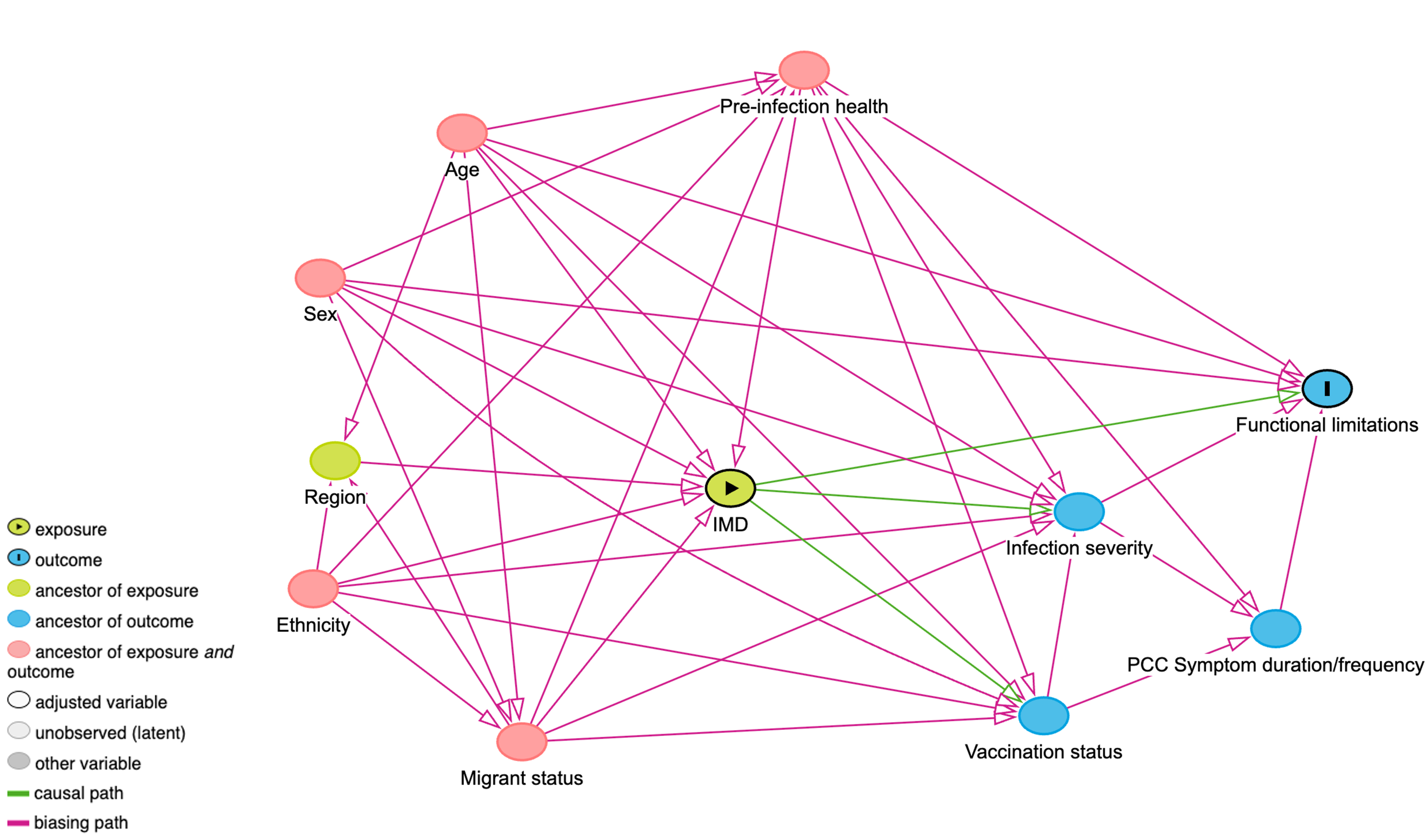

**Figure 2 Directed Acyclic Graph for the impact of deprivation (IMD Quintile) on experiencing functional limitations among people with PCC. This adjustment set holds the assumptions of pre-infection health influencing deprivation.**

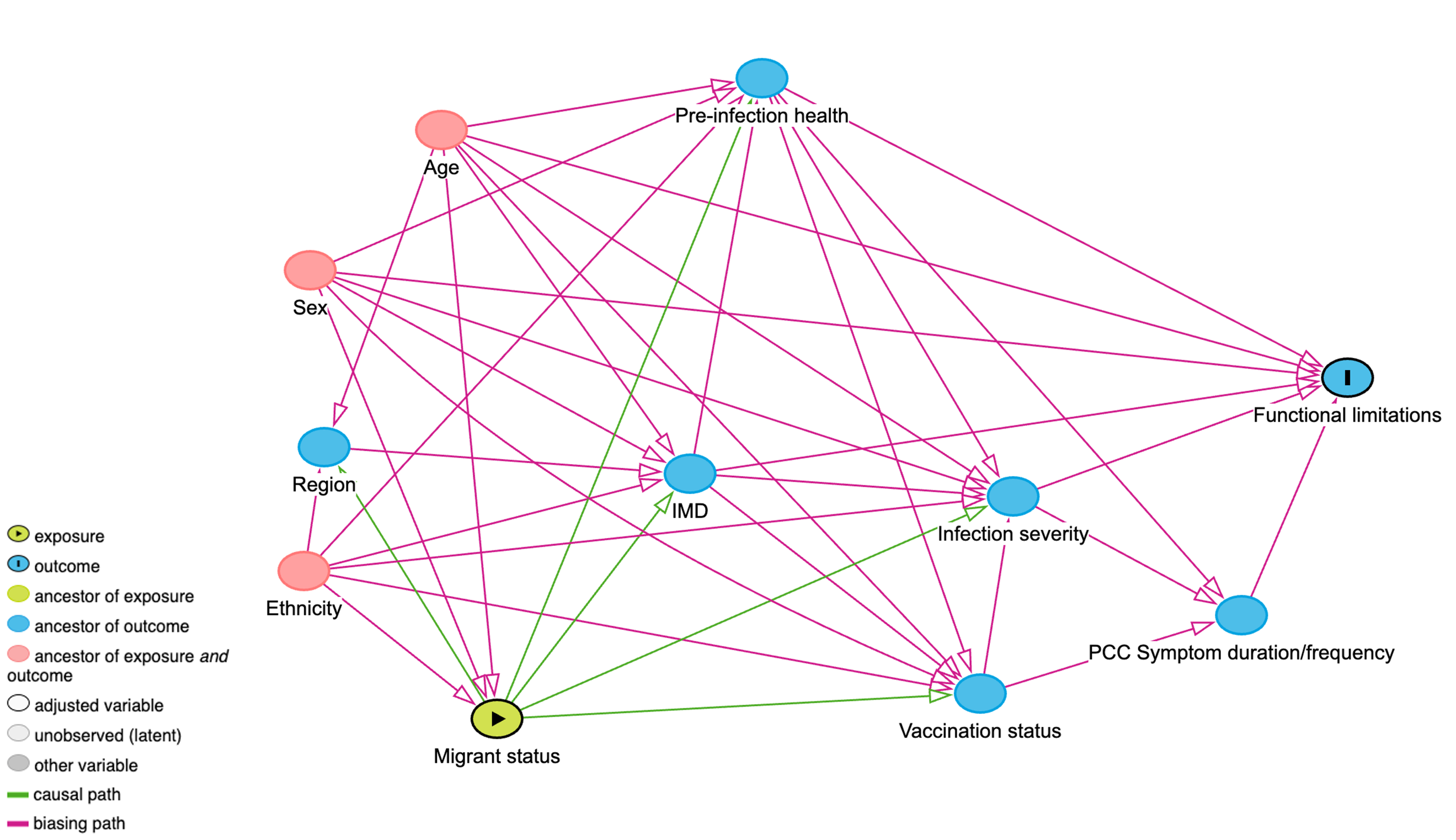

**Figure 3 Directed Acyclic Graph for the impact of migration status on experiencing functional limitations among people with PCC. This adjustment set holds the assumptions of migration status quintile influencing pre-infection health.**

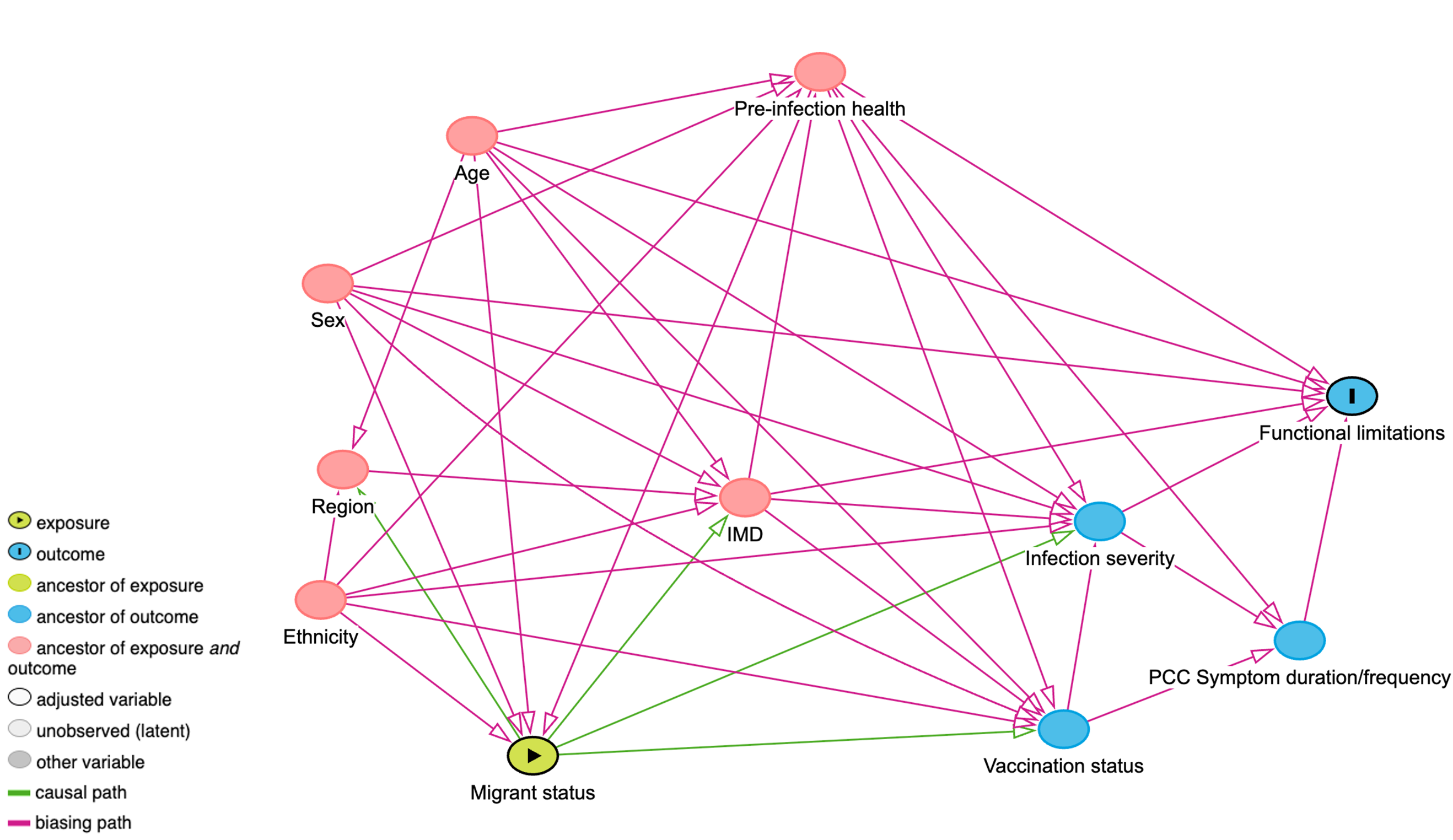

**Figure 4 Directed Acyclic Graph for the impact of migration status on experiencing functional limitations among people with PCC. This adjustment set holds the assumptions of migration status influencing pre-infection health.**

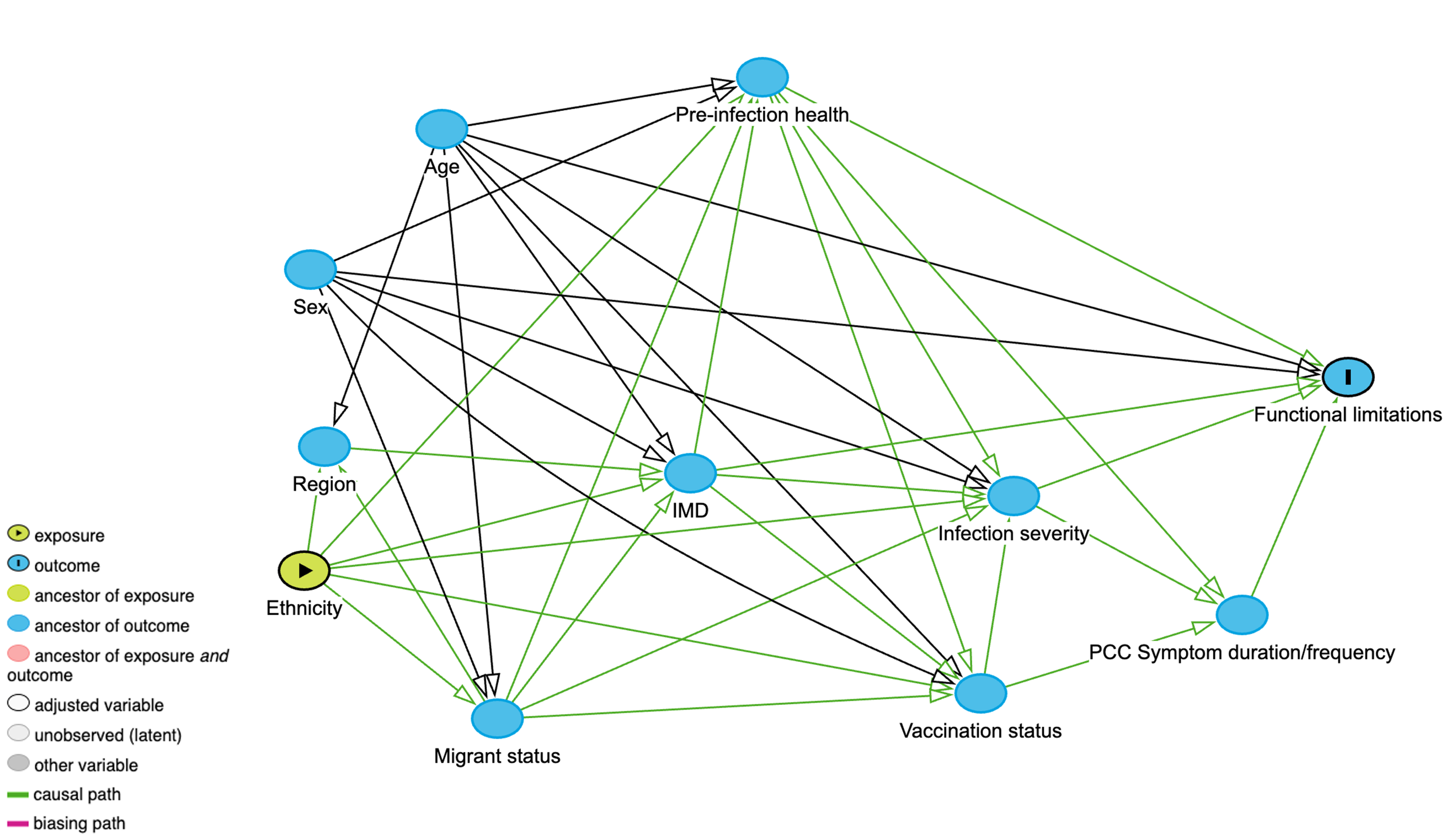

**Figure 5 Directed Acyclic Graph for the impact of ethnicity on experiencing functional limitations among people with PCC.**

**Table 1 Sociodemographic and clinical characteristics of the analysis cohorts by functional limitation.**

|  | **Attend or participate in work or education** | **Concentrate on things** | **Take care of yourself** | **Take care of others in the household** | **Do necessary daily activities outside the household** | **Do activities that you enjoy** |
| --- | --- | --- | --- | --- | --- | --- |
| **Characteristic** | **N = 530** | **N = 743** | **N = 713** | **N = 593** | **N = 740** | **N = 748** |
| **Age group** |  |  |  |  |  |  |
| 18-44 | 92 (17%) | 91 (12%) | 92 (13%) | 80 (13%) | 91 (12%) | 91 (12%) |
| 45-64 | 304 (57%) | 368 (50%) | 351 (49%) | 300 (51%) | 362 (49%) | 364 (49%) |
| 65+ | 134 (25%) | 284 (38%) | 270 (38%) | 213 (36%) | 287 (39%) | 293 (39%) |
| **Sex (at birth)** |  |  |  |  |  |  |
| Male | 152 (29%) | 211 (28%) | 206 (29%) | 176 (30%) | 212 (29%) | 217 (29%) |
| Female | 378 (71%) | 532 (72%) | 507 (71%) | 417 (70%) | 528 (71%) | 531 (71%) |
| **IMD Quintile** |  |  |  |  |  |  |
| 1 | 52 (9.8%) | 65 (8.7%) | 59 (8.3%) | 51 (8.6%) | 66 (8.9%) | 63 (8.4%) |
| 2 | 107 (20%) | 134 (18%) | 129 (18%) | 100 (17%) | 131 (18%) | 135 (18%) |
| 3 | 116 (22%) | 158 (21%) | 154 (22%) | 121 (20%) | 158 (21%) | 160 (21%) |
| 4 | 122 (23%) | 180 (24%) | 175 (25%) | 139 (23%) | 183 (25%) | 182 (24%) |
| 5 | 125 (24%) | 197 (27%) | 187 (26%) | 174 (29%) | 193 (26%) | 199 (27%) |
| Missing | 8 (1.5%) | 9 (1.2%) | 9 (1.3%) | 8 (1.3%) | 9 (1.2%) | 9 (1.2%) |
| **IMD Quintile (excluding the health domain)** |  |  |  |  |  |  |
| 1 | 50 (9.4%) | 65 (8.7%) | 59 (8.3%) | 50 (8.4%) | 64 (8.6%) | 63 (8.4%) |
| 2 | 108 (20%) | 134 (18%) | 129 (18%) | 100 (17%) | 132 (18%) | 135 (18%) |
| 3 | 129 (24%) | 176 (24%) | 171 (24%) | 134 (23%) | 177 (24%) | 175 (23%) |
| 4 | 119 (22%) | 182 (24%) | 179 (25%) | 146 (25%) | 186 (25%) | 188 (25%) |
| 5 | 116 (22%) | 177 (24%) | 166 (23%) | 155 (26%) | 172 (23%) | 178 (24%) |
| Missing | 8 (1.5%) | 9 (1.2%) | 9 (1.3%) | 8 (1.3%) | 9 (1.2%) | 9 (1.2%) |
| **Migration status** |  |  |  |  |  |  |
| UK Born | 352 (66%) | 514 (69%) | 487 (68%) | 409 (69%) | 512 (69%) | 518 (69%) |
| Not UK-born | 48 (9.1%) | 54 (7.3%) | 56 (7.9%) | 44 (7.4%) | 55 (7.4%) | 56 (7.5%) |
| Missing | 130 (25%) | 175 (24%) | 170 (24%) | 140 (24%) | 173 (23%) | 174 (23%) |
| **Ethnic minority status** |  |  |  |  |  |  |
| White British | 451 (85%) | 655 (88%) | 623 (87%) | 522 (88%) | 650 (88%) | 658 (88%) |
| Ethnic minority | 69 (13%) | 77 (10%) | 79 (11%) | 64 (11%) | 79 (11%) | 79 (11%) |
| Missing/Prefer not to say | 10 (1.9%) | 11 (1.5%) | 11 (1.5%) | 7 (1.2%) | 11 (1.5%) | 11 (1.5%) |
| **Clinical vulnerability** |  |  |  |  |  |  |
| Not clinically vulnerable | 270 (51%) | 359 (48%) | 342 (48%) | 292 (49%) | 358 (48%) | 361 (48%) |
| Clinically vulnerable | 160 (30%) | 243 (33%) | 233 (33%) | 186 (31%) | 244 (33%) | 243 (32%) |
| Clinically extremely vulnerable | 82 (15%) | 123 (17%) | 120 (17%) | 99 (17%) | 120 (16%) | 126 (17%) |
| Missing | 18 (3.4%) | 18 (2.4%) | 18 (2.5%) | 16 (2.7%) | 18 (2.4%) | 18 (2.4%) |

### **Multicollinearity**

We produced generated the adjusted generalised standard error inflation factor (aGSIF) to detect multicollinearity in models. No evidence of multicollinearity was detected, with all aGSIF values below 2, respectively (see below Tables ).

**Table 2 Adjusted generalised standard error inflation factor (aGSIF) for regression models representing the total effect of IMD quintile on experiencing functional limitations.** aGSIF of below 2 suggest a lack of evidence of multicollinearity.

| **Exposure: IMD Quintile** | **aGSIF** | | | | | |
| --- | --- | --- | --- | --- | --- | --- |
| **Analysis cohort** | Attending or participating in work/education | Concentrating | Self-care | Taking care of others in the household | Doing necessary activities outside the household | Doing enjoyable activities |
| IMD Quintile | 1.01 | 1.01 | 1.01 | 1.01 | 1.01 | 1.01 |
| Age group | 1.02 | 1.02 | 1.03 | 1.02 | 1.02 | 1.02 |
| Sex-at-birth | 1.01 | 1.01 | 1.01 | 1.01 | 1.01 | 1.01 |
| Minority ethnicity status | 1.26 | 1.27 | 1.33 | 1.27 | 1.31 | 1.29 |
| Migration status | 1.11 | 1.12 | 1.14 | 1.12 | 1.14 | 1.13 |

**Table 3 Adjusted generalised standard error inflation factor (aGSIF) for regression models representing the total effect of migration status on experiencing functional limitations.** aGSIF of below 2 suggest a lack of evidence of multicollinearity.

| **Exposure: Migration Status** | **aGSIF** | | | | | |
| --- | --- | --- | --- | --- | --- | --- |
| **Analysis cohort** | Attending or participating in work/education | Concentrating | Self-care | Taking care of others in the household | Doing necessary activities outside the household | Doing enjoyable activities |
| Migration status | 1.33 | 1.37 | 1.41 | 1.37 | 1.40 | 1.37 |
| Age group | 1.01 | 1.01 | 1.02 | 1.01 | 1.02 | 1.02 |
| Sex-at-birth | 1.00 | 1.00 | 1.01 | 1.00 | 1.01 | 1.01 |
| Minority ethnicity status | 1.34 | 1.38 | 1.43 | 1.38 | 1.42 | 1.39 |

**Table 4** **Adjusted generalised standard error inflation factor (aGSIF) for regression models representing the total effect of minority ethnicity status on experiencing functional limitations.** aGSIF of below 2 suggest a lack of evidence of multicollinearity.

| **Exposure: Ethnic Minority Status** | **aGSIF** | | | | | |
| --- | --- | --- | --- | --- | --- | --- |
| **Analysis cohort** | Attending or participating in work/education | Concentrating | Self-care | Taking care of others in the household | Doing necessary activities outside the household | Doing enjoyable activities |
| Minority ethnicity status | 1.04 | 1.03 | 1.04 | 1.03 | 1.04 | 1.04 |
| Age group | 1.02 | 1.02 | 1.02 | 1.01 | 1.02 | 1.02 |
| Sex-at-birth | 1.00 | 1.00 | 1.01 | 1.00 | 1.00 | 1.01 |

**Sociodemographic and clinical characteristics by exposure and outcome**

**Main Analyses - Unadjusted and Adjusted Odds Ratios**

**Sensitivity Analyses - Unadjusted and Adjusted Odds Ratios**

**Main Analysis – Odds Ratios (Exposure: IMD Quintile)**

**Table 5 Unadjusted odds ratio (OR) and adjusted odds ratio (aOR) for the association between IMD quintile and experiencing limitations in attending or participating in work or education.**

|  | Unadjusted |  | Adjusted |  | Adjusted + |  |
| --- | --- | --- | --- | --- | --- | --- |
|  | **OR** | **95% CI** | **aOR** | **95% CI** | **aOR** | **95% CI** |
| **IMD Quintile** |  |  |  |  |  |  |
| 1 (most deprived) | 2.68 | 1.33, 5.67 | 2.46 | 1.21, 5.24 | 2.31 | 1.13, 4.95 |
| 2 | 1.95 | 1.14, 3.37 | 1.88 | 1.09, 3.29 | 1.96 | 1.12, 3.47 |
| 3 | 1.27 | 0.76, 2.13 | 1.23 | 0.73, 2.08 | 1.22 | 0.72, 2.08 |
| 4 | 1.24 | 0.75, 2.06 | 1.28 | 0.76, 2.15 | 1.27 | 0.75, 2.15 |
| 5 (least deprived) | ref | - | ref | - | ref | - |
| **Age group** |  |  |  |  |  |  |
| 18-44 |  |  | ref | - | ref | - |
| 45-64 |  |  | 0.93 | 0.55, 1.56 | 0.95 | 0.56, 1.60 |
| 65+ |  |  | 0.52 | 0.29, 0.93 | 0.52 | 0.29, 0.94 |
| **Sex-at-birth** |  |  |  |  |  |  |
| Male |  |  | ref | - | ref | - |
| Female |  |  | 0.98 | 0.65, 1.46 | 0.95 | 0.63, 1.43 |
| **Minority ethnicity status** |  |  |  |  |  |  |
| White British |  |  | ref | - | ref | - |
| Ethnic minority |  |  | 0.80 | 0.40, 1.60 | 0.75 | 0.37, 1.50 |
| **Migration status** |  |  |  |  |  |  |
| UK Born |  |  | ref | - | ref | - |
| Not UK Born |  |  | 1.58 | 0.70, 3.68 | 1.61 | 0.71, 3.80 |
| Missing |  |  | 0.93 | 0.60, 1.43 | 0.95 | 0.60, 1.49 |
| **Clinical vulnerability** |  |  |  |  |  |  |
| Not clinically vulnerable |  |  |  |  | ref | - |
| Clinically vulnerable |  |  |  |  | 1.37 | 0.90, 2.08 |
| Clinically extremely vulnerable |  |  |  |  | 1.17 | 0.69, 2.01 |

**Table 6 Unadjusted odds ratio (OR) and adjusted odds ratio (aOR) for the association between IMD quintile and experiencing limitations in concentrating.**

|  | Unadjusted |  | Adj 1 |  | Adj + CV |  |
| --- | --- | --- | --- | --- | --- | --- |
|  | **OR** | **95% CI** | **aOR** | **95% CI** | **aOR** | **95% CI** |
| **IMD Quintile** |  |  |  |  |  |  |
| 1 (most deprived) | 3.03 | 1.57, 6.29 | 2.85 | 1.46, 5.93 | 2.76 | 1.41, 5.76 |
| 2 | 1.90 | 1.18, 3.10 | 1.85 | 1.15, 3.03 | 1.92 | 1.18, 3.16 |
| 3 | 1.50 | 0.97, 2.35 | 1.47 | 0.94, 2.30 | 1.46 | 0.93, 2.29 |
| 4 | 1.13 | 0.75, 1.72 | 1.15 | 0.75, 1.76 | 1.16 | 0.76, 1.77 |
| 5 (least deprived) | ref | - | ref | - | ref | - |
| **Age group** |  |  |  |  |  |  |
| 18-44 |  |  | ref | - | ref | - |
| 45-64 |  |  | 0.96 | 0.55, 1.63 | 1.02 | 0.58, 1.74 |
| 65+ |  |  | 0.68 | 0.39, 1.17 | 0.72 | 0.41, 1.24 |
| **Sex-at-birth** |  |  |  |  |  |  |
| Male |  |  | ref | - | ref | - |
| Female |  |  | 1.13 | 0.80, 1.60 | 1.17 | 0.82, 1.65 |
| **Minority ethnicity status** |  |  |  |  |  |  |
| White British |  |  | ref | - | ref | - |
| Ethnic minority |  |  | 1.46 | 0.74, 3.03 | 1.44 | 0.73, 2.99 |
| **Migration status** |  |  |  |  |  |  |
| UK Born |  |  | ref | - | ref | - |
| Not UK Born |  |  | 0.88 | 0.40, 1.99 | 0.87 | 0.39, 1.98 |
| Missing |  |  | 1.23 | 0.84, 1.82 | 1.21 | 0.82, 1.82 |
| **Clinical vulnerability** |  |  |  |  |  |  |
| Not clinically vulnerable |  |  |  |  | ref | - |
| Clinically vulnerable |  |  |  |  | 1.15 | 0.80, 1.64 |
| Clinically extremely vulnerable |  |  |  |  | 1.20 | 0.76, 1.90 |

**Table 7 Unadjusted odds ratio (OR) and adjusted odds ratio (aOR) for the association between IMD quintile and experiencing limitations in self-care.**

| Impact 3 | Unadjusted |  | Adj 1 |  | Adj + CV |  |
| --- | --- | --- | --- | --- | --- | --- |
|  | **OR** | **95% CI** | **aOR** | **95% CI** | **aOR** | **95% CI** |
| **IMD Quintile** |  |  |  |  |  |  |
| 1 (most deprived) | 2.53 | 1.31, 4.85 | 2.42 | 1.24, 4.69 | 2.04 | 1.02, 4.04 |
| 2 | 1.67 | 0.97, 2.89 | 1.61 | 0.93, 2.80 | 1.68 | 0.96, 2.94 |
| 3 | 1.69 | 1.01, 2.86 | 1.70 | 1.01, 2.88 | 1.67 | 0.99, 2.86 |
| 4 | 1.88 | 1.15, 3.12 | 2.01 | 1.22, 3.36 | 2.03 | 1.22, 3.40 |
| 5 (least deprived) | ref | - | ref | - | ref | - |
| **Age group** |  |  |  |  |  |  |
| 18-44 |  |  | ref | - | ref | - |
| 45-64 |  |  | 0.56 | 0.33, 0.94 | 0.52 | 0.31, 0.89 |
| 65+ |  |  | 0.64 | 0.37, 1.11 | 0.58 | 0.33, 1.01 |
| **Sex-at-birth** |  |  |  |  |  |  |
| Male |  |  | ref | - | ref | - |
| Female |  |  | 0.92 | 0.63, 1.36 | 0.95 | 0.65, 1.41 |
| **Minority ethnicity status** |  |  |  |  |  |  |
| White British |  |  | ref | - | ref | - |
| Ethnic minority |  |  | 1.19 | 0.59, 2.33 | 1.23 | 0.60, 2.45 |
| **Migration status** |  |  |  |  |  |  |
| UK Born |  |  | ref | - | ref | - |
| Not UK Born |  |  | 1.26 | 0.57, 2.72 | 1.22 | 0.54, 2.69 |
| Missing |  |  | 1.00 | 0.65, 1.52 | 1.12 | 0.72, 1.73 |
| **Clinical vulnerability** |  |  |  |  |  |  |
| Not clinically vulnerable |  |  |  |  | ref | - |
| Clinically vulnerable |  |  |  |  | 1.53 | 1.03, 2.26 |
| Clinically extremely vulnerable |  |  |  |  | 1.91 | 1.18, 3.07 |

**Table 8 Unadjusted odds ratio (OR) and adjusted odds ratio (aOR) for the association between IMD quintile and experiencing limitations in taking care of others in the household.**

|  | Unadjusted |  | Adj 1 |  | Adj + CV |  |
| --- | --- | --- | --- | --- | --- | --- |
|  | **OR** | **95% CI** | **aOR** | **95% CI** | **aOR** | **95% CI** |
| **IMD Quintile** |  |  |  |  |  |  |
| 1 (most deprived) | 1.96 | 1.03, 3.71 | 1.90 | 0.99, 3.62 | 1.75 | 0.90, 3.37 |
| 2 | 1.15 | 0.68, 1.94 | 1.11 | 0.65, 1.88 | 1.11 | 0.64, 1.90 |
| 3 | 0.88 | 0.53, 1.47 | 0.86 | 0.51, 1.44 | 0.86 | 0.51, 1.44 |
| 4 | 1.20 | 0.75, 1.93 | 1.15 | 0.71, 1.86 | 1.14 | 0.70, 1.85 |
| 5 (least deprived) | ref | - | ref | - | ref | - |
| **Age group** |  |  |  |  |  |  |
| 18-44 |  |  | ref | - | ref | - |
| 45-64 |  |  | 0.65 | 0.38, 1.11 | 0.59 | 0.34, 1.01 |
| 65+ |  |  | 0.65 | 0.37, 1.14 | 0.6 | 0.34, 1.06 |
| **Sex-at-birth** |  |  |  |  |  |  |
| Male |  |  | ref | - | ref | - |
| Female |  |  | 1.19 | 0.81, 1.76 | 1.26 | 0.85, 1.89 |
| **Minority ethnicity status** |  |  |  |  |  |  |
| White British |  |  | ref | - | ref | - |
| Ethnic minority |  |  | 0.76 | 0.35, 1.56 | 0.80 | 0.37, 1.66 |
| **Migration status** |  |  |  |  |  |  |
| UK Born |  |  | ref | - | ref | - |
| Not UK Born |  |  | 0.79 | 0.32, 1.89 | 0.76 | 0.30, 1.84 |
| Missing |  |  | 0.91 | 0.59, 1.39 | 0.97 | 0.62, 1.50 |
| **Clinical vulnerability** |  |  |  |  |  |  |
| Not clinically vulnerable |  |  |  |  | ref | - |
| Clinically vulnerable |  |  |  |  | 1.45 | 0.98, 2.16 |
| Clinically extremely vulnerable |  |  |  |  | 1.50 | 0.91, 2.44 |

**Table 9 Unadjusted odds ratio (OR) and adjusted odds ratio (aOR) for the association between IMD quintile and experiencing limitations in doing necessary activities outside the house.**

|  | Unadjusted |  | Adj 1 |  | Adj + CV |  |
| --- | --- | --- | --- | --- | --- | --- |
|  | **OR** | **95% CI** | **aOR** | **95% CI** | **aOR** | **95% CI** |
| **IMD Quintile** |  |  |  |  |  |  |
| 1 (most deprived) | 2.21 | 1.21, 4.15 | 2.20 | 1.20, 4.15 | 2.04 | 1.10, 3.91 |
| 2 | 1.40 | 0.89, 2.21 | 1.38 | 0.87, 2.19 | 1.42 | 0.89, 2.28 |
| 3 | 1.10 | 0.72, 1.69 | 1.07 | 0.70, 1.65 | 1.09 | 0.70, 1.69 |
| 4 | 1.43 | 0.95, 2.17 | 1.38 | 0.91, 2.10 | 1.39 | 0.90, 2.13 |
| 5 (least deprived) | ref | - | ref | - | ref | - |
| **Age group** |  |  |  |  |  |  |
| 18-44 |  |  | ref | - | ref | - |
| 45-64 |  |  | 0.9 | 0.54, 1.47 | 0.84 | 0.50, 1.4 |
| 65+ |  |  | 1.00 | 0.59, 1.67 | 0.91 | 0.53, 1.54 |
| **Sex-at-birth** |  |  |  |  |  |  |
| Male |  |  | ref | - | ref | - |
| Female |  |  | 1.36 | 0.98, 1.90 | 1.41 | 1.01, 1.98 |
| **Minority ethnicity status** |  |  |  |  |  |  |
| White British |  |  | ref | - | ref | - |
| Ethnic minority |  |  | 0.70 | 0.37, 1.32 | 0.70 | 0.37, 1.34 |
| **Migration status** |  |  |  |  |  |  |
| UK Born |  |  | ref | - | ref | - |
| Not UK Born |  |  | 1.32 | 0.63, 2.82 | 1.21 | 0.57, 2.62 |
| Missing |  |  | 0.92 | 0.64, 1.33 | 1.06 | 0.73, 1.57 |
| **Clinical vulnerability** |  |  |  |  |  |  |
| Not clinically vulnerable |  |  |  |  | ref | - |
| Clinically vulnerable |  |  |  |  | 1.79 | 1.27, 2.53 |
| Clinically extremely vulnerable |  |  |  |  | 2.02 | 1.29, 3.22 |

**Table 10 Unadjusted odds ratio (OR) and adjusted odds ratio (aOR) for the association between IMD quintile and experiencing limitations in doing enjoyable activities.**

|  | Unadjusted |  | Adj 1 |  | Adj + CV |  |
| --- | --- | --- | --- | --- | --- | --- |
|  | **OR** | **95% CI** | **aOR** | **95% CI** | **aOR** | **95% CI** |
| **IMD Quintile** |  |  |  |  |  |  |
| 1 (most deprived) | 1.88 | 0.96, 3.91 | 1.86 | 0.94, 3.89 | 1.78 | 0.90, 3.74 |
| 2 | 1.10 | 0.68, 1.80 | 1.09 | 0.67, 1.78 | 1.11 | 0.68, 1.83 |
| 3 | 1.07 | 0.68, 1.70 | 1.06 | 0.67, 1.69 | 1.07 | 0.67, 1.71 |
| 4 | 1.38 | 0.88, 2.19 | 1.37 | 0.87, 2.17 | 1.36 | 0.86, 2.18 |
| 5 (least deprived) | ref | - | ref | - | ref | - |
| **Age group** |  |  |  |  |  |  |
| 18-44 |  |  | ref | - | ref | - |
| 45-64 |  |  | 0.81 | 0.46, 1.40 | 0.82 | 0.46, 1.42 |
| 65+ |  |  | 0.93 | 0.51, 1.63 | 0.91 | 0.50, 1.61 |
| **Sex-at-birth** |  |  |  |  |  |  |
| Male |  |  | ref | - | ref | - |
| Female |  |  | 1.20 | 0.84, 1.71 | 1.19 | 0.83, 1.71 |
| **Minority ethnicity status** |  |  |  |  |  |  |
| White British |  |  | ref | - | ref | - |
| Ethnic minority |  |  | 0.82 | 0.42, 1.65 | 0.77 | 0.39, 1.56 |
| **Migration status** |  |  |  |  |  |  |
| UK Born |  |  | ref | - | ref | - |
| Not UK Born |  |  | 1.30 | 0.59, 2.98 | 1.31 | 0.59, 3.02 |
| Missing |  |  | 1.00 | 0.67, 1.50 | 1.08 | 0.71, 1.64 |
| **Clinical vulnerability** |  |  |  |  |  |  |
| Not clinically vulnerable |  |  |  |  | ref | - |
| Clinically vulnerable |  |  |  |  | 1.37 | 0.95, 2.01 |
| Clinically extremely vulnerable |  |  |  |  | 1.27 | 0.80, 2.06 |

**Main Analysis – Odds Ratios (Exposure: Migration Status)**

**Table 11 Unadjusted odds ratio (OR) and adjusted odds ratio (aOR) for the association between migration status and experiencing limitations in attending or participating in work or education.**

|  | Unadjusted |  | Adj 1 |  | Adj + CV |  |
| --- | --- | --- | --- | --- | --- | --- |
|  | **OR** | **95% CI** | **aOR** | **95% CI** | **aOR** | **95% CI** |
| **Migration Status** |  |  |  |  |  |  |
| UK Born | ref | - | ref | - | ref | - |
| Not UK Born | 1.55 | 0.82, 3.08 | 1.55 | 0.65, 3.85 | 1.49 | 0.62, 3.73 |
| **Age group** |  |  |  |  |  |  |
| 18-44 |  |  | ref | - | ref | - |
| 45-64 |  |  | 0.93 | 0.52, 1.63 | 0.94 | 0.52, 1.65 |
| 65+ |  |  | 0.43 | 0.22, 0.82 | 0.43 | 0.22, 0.83 |
| **Sex-at-birth** |  |  |  |  |  |  |
| Male |  |  | ref | - | ref | - |
| Female |  |  | 1.01 | 0.64, 1.58 | 1.01 | 0.64, 1.58 |
| **Minority Ethnicity Status** |  |  |  |  |  |  |
| White British |  |  | ref | - | ref | - |
| Ethnic minority |  |  | 0.81 | 0.36, 1.84 | 0.81 | 0.36, 1.83 |
| **Clinical vulnerability** |  |  |  |  |  |  |
| Not clinically vulnerable |  |  |  |  | ref | - |
| Clinically vulnerable |  |  |  |  | 1.20 | 0.76, 1.92 |
| Clinically extremely vulnerable |  |  |  |  | 1.20 | 0.68, 2.15 |

**Table 12 Unadjusted odds ratio (OR) and adjusted odds ratio (aOR) for the association between migration status and experiencing limitations in concentrating.**

|  | Unadjusted |  | Adj 1 |  | Adj + CV |  |
| --- | --- | --- | --- | --- | --- | --- |
|  | **OR** | **95% CI** | **aOR** | **95% CI** | **aOR** | **95% CI** |
| **Migration Status** |  |  |  |  |  |  |
| UK Born | ref | - | ref | - | ref | - |
| Not UK Born | 1.39 | 0.76, 2.66 | 0.69 | 0.29, 1.66 | 0.66 | 0.27, 1.59 |
| **Age group** |  |  |  |  |  |  |
| 18-44 |  |  | ref | - | ref | - |
| 45-64 |  |  | 1.07 | 0.59, 1.89 | 1.08 | 0.60, 1.91 |
| 65+ |  |  | 0.67 | 0.36, 1.20 | 0.67 | 0.37, 1.21 |
| **Sex-at-birth** |  |  |  |  |  |  |
| Male |  |  | ref | - | ref | - |
| Female |  |  | 1.22 | 0.83, 1.79 | 1.22 | 0.83, 1.79 |
| **Minority Ethnicity Status** |  |  |  |  |  |  |
| White British |  |  | ref | - | ref | - |
| Ethnic minority |  |  | 2.37 | 1.01, 6.04 | 2.38 | 1.02, 6.07 |
| **Clinical vulnerability** |  |  |  |  |  |  |
| Not clinically vulnerable |  |  |  |  | ref | - |
| Clinically vulnerable |  |  |  |  | 0.99 | 0.67, 1.48 |
| Clinically extremely vulnerable |  |  |  |  | 1.19 | 0.74, 1.94 |

**Table 13 Unadjusted odds ratio (OR) and adjusted odds ratio (aOR) for the association between migration status and experiencing limitations in self-care.**

|  | Unadjusted |  | Adj 1 |  | Adj + CV |  |
| --- | --- | --- | --- | --- | --- | --- |
|  | **OR** | **95% CI** | **aOR** | **95% CI** | **aOR** | **95% CI** |
| **Migration Status** |  |  |  |  |  |  |
| UK Born | ref | - | ref | - | ref | - |
| Not UK Born | 1.52 | 0.83, 2.71 | 1.10 | 0.46, 2.54 | 1.03 | 0.43, 2.41 |
| **Age group** |  |  |  |  |  |  |
| 18-44 |  |  | ref | - | ref | - |
| 45-64 |  |  | 0.51 | 0.30, 0.90 | 0.50 | 0.29, 0.88 |
| 65+ |  |  | 0.50 | 0.28, 0.91 | 0.48 | 0.26, 0.87 |
| **Sex-at-birth** |  |  |  |  |  |  |
| Male |  |  | ref | - | ref | - |
| Female |  |  | 0.87 | 0.57, 1.33 | 0.89 | 0.58, 1.37 |
| **Minority Ethnicity Status** |  |  |  |  |  |  |
| White British |  |  | ref | - | ref | - |
| Ethnic minority |  |  | 1.31 | 0.57, 2.90 | 1.40 | 0.61, 3.14 |
| **Clinical vulnerability** |  |  |  |  |  |  |
| Not clinically vulnerable |  |  |  |  | ref | - |
| Clinically vulnerable |  |  |  |  | 1.10 | 0.70, 1.73 |
| Clinically extremely vulnerable |  |  |  |  | 1.77 | 1.07, 2.92 |

**Table 14 Unadjusted odds ratio (OR) and adjusted odds ratio (aOR) for the association between migration status and experiencing limitations in taking care of others in the household.**

|  | Unadjusted |  | Adj 1 |  | Adj + CV |  |
| --- | --- | --- | --- | --- | --- | --- |
|  | **OR** | **95% CI** | **aOR** | **95% CI** | **aOR** | **95% CI** |
| **Migration Status** |  |  |  |  |  |  |
| UK Born | ref | - | ref | - | ref | - |
| Not UK Born | 0.71 | 0.34, 1.38 | 0.65 | 0.24, 1.68 | 0.65 | 0.24, 1.68 |
| **Age group** |  |  |  |  |  |  |
| 18-44 |  |  | ref | - | ref | - |
| 45-64 |  |  | 0.58 | 0.32, 1.03 | 0.56 | 0.32, 1.01 |
| 65+ |  |  | 0.58 | 0.32, 1.08 | 0.56 | 0.30, 1.04 |
| **Sex-at-birth** |  |  |  |  |  |  |
| Male |  |  | ref | - | ref | - |
| Female |  |  | 1.23 | 0.80, 1.91 | 1.27 | 0.83, 1.98 |
| **Minority Ethnicity Status** |  |  |  |  |  |  |
| White British |  |  | ref | - | ref | - |
| Ethnic minority |  |  | 0.94 | 0.38, 2.28 | 0.97 | 0.39, 2.35 |
| **Clinical vulnerability** |  |  |  |  |  |  |
| Not clinically vulnerable |  |  |  |  | ref | - |
| Clinically vulnerable |  |  |  |  | 1.30 | 0.83, 2.03 |
| Clinically extremely vulnerable |  |  |  |  | 1.31 | 0.77, 2.21 |

**Table 15 Unadjusted odds ratio (OR) and adjusted odds ratio (aOR) for the association between migration status and experiencing limitations in doing necessary activities outside the house.**

|  | Unadjusted |  | Adj 1 |  | Adj + CV |  |
| --- | --- | --- | --- | --- | --- | --- |
|  | **OR** | **95% CI** | **aOR** | **95% CI** | **aOR** | **95% CI** |
| **Migration Status** |  |  |  |  |  |  |
| UK Born | ref | - | ref | - |  |  |
| Not UK Born | 1.03 | 0.59, 1.85 | 1.27 | 0.57, 2.90 | ref | - |
| **Age group** |  |  |  |  | 1.16 | 0.51, 2.66 |
| 18-44 |  |  | ref | - |  |  |
| 45-64 |  |  | 0.83 | 0.47, 1.43 | ref | - |
| 65+ |  |  | 0.93 | 0.52, 1.64 | 0.86 | 0.49, 1.48 |
| **Sex-at-birth** |  |  |  |  | 0.91 | 0.51, 1.61 |
| Male |  |  | ref | - |  |  |
| Female |  |  | 1.47 | 1.02, 2.13 | ref | - |
| **Minority Ethnicity Status** |  |  |  |  | 1.53 | 1.05, 2.23 |
| White British |  |  | ref | - |  |  |
| Ethnic minority |  |  | 0.72 | 0.34, 1.55 | ref | - |
| **Clinical vulnerability** |  |  |  |  | 0.73 | 0.34, 1.57 |
| Not clinically vulnerable |  |  |  |  |  |  |
| Clinically vulnerable |  |  |  |  | ref | - |
| Clinically extremely vulnerable |  |  |  |  | 1.57 | 1.07, 2.32 |

**Table 16 Unadjusted odds ratio (OR) and adjusted odds ratio (aOR) for the association between migration status and experiencing limitations in doing enjoyable activities.**

|  | Unadjusted |  | Adj 1 |  | Adj + CV |  |
| --- | --- | --- | --- | --- | --- | --- |
|  | **OR** | **95% CI** | **aOR** | **95% CI** | **aOR** | **95% CI** |
| **Migration Status** |  |  |  |  |  |  |
| UK Born | ref | - | ref | - | ref | - |
| Not UK Born | 1.13 | 0.61, 2.21 | 1.26 | 0.54, 3.10 | 1.21 | 0.51, 2.97 |
| **Age group** |  |  |  |  |  |  |
| 18-44 |  |  | ref | - | ref | - |
| 45-64 |  |  | 0.82 | 0.44, 1.47 | 0.83 | 0.44, 1.49 |
| 65+ |  |  | 0.99 | 0.52, 1.84 | 0.99 | 0.52, 1.85 |
| **Sex-at-birth** |  |  |  |  |  |  |
| Male |  |  | ref | - | ref | - |
| Female |  |  | 1.07 | 0.71, 1.59 | 1.07 | 0.71, 1.60 |
| **Minority Ethnicity Status** |  |  |  |  |  |  |
| White British |  |  | ref | - | ref | - |
| Ethnic minority |  |  | 0.86 | 0.39, 2.00 | 0.87 | 0.39, 2.02 |
| **Clinical vulnerability** |  |  |  |  |  |  |
| Not clinically vulnerable |  |  |  |  | ref | - |
| Clinically vulnerable |  |  |  |  | 1.09 | 0.72, 1.66 |
| Clinically extremely vulnerable |  |  |  |  | 1.27 | 0.77, 2.14 |

**Main Analyses – Odds Ratios (Exposure: Ethnic minority status)**

**Table 17 Unadjusted odds ratio (OR) and adjusted odds ratio (aOR) for the association between ethnic minority status and experiencing limitations in attending or participating in work or education.**

|  | **Unadjusted model** | | **Adjusted model** | |
| --- | --- | --- | --- | --- |
|  | **OR** | **95% CI** | **aOR** | **95% CI** |
| **Minority ethnicity status** |  |  |  |  |
| White British | ref | - | ref | - |
| Ethnic minority | 1.14 | 0.68, 1.95 | 1.00 | 0.58, 1.76 |
| **Age group** |  |  |  |  |
| 18-44 |  |  | ref | - |
| 45-64 |  |  | 0.91 | 0.54, 1.51 |
| 65+ |  |  | 0.47 | 0.26, 0.83 |
| **Sex-at-birth** |  |  |  |  |
| Male |  |  | ref | - |
| Female |  |  | 1.00 | 0.67, 1.48 |

**Table 18 Unadjusted odds ratio (OR) and adjusted odds ratio (aOR) for the association between ethnic minority status and experiencing limitations in concentrating.**

|  | **Unadjusted model** | | **Adjusted model** | |
| --- | --- | --- | --- | --- |
|  | **OR** | **95% CI** | **aOR** | **95% CI** |
| **Minority ethnicity status** |  |  |  |  |
| White British | ref | - | ref | - |
| Ethnic minority | 1.60 | 0.95, 2.81 | 1.41 | 0.82, 2.53 |
| **Age group** |  |  |  |  |
| 18-44 |  |  | ref | - |
| 45-64 |  |  | 0.92 | 0.53, 1.54 |
| 65+ |  |  | 0.61 | 0.35, 1.03 |
| **Sex-at-birth** |  |  |  |  |
| Male |  |  | ref | - |
| Female |  |  | 1.13 | 0.81, 1.59 |

**Table 19 Unadjusted odds ratio (OR) and adjusted odds ratio (aOR) for the association between ethnic minority status and experiencing limitations in self-care.**

|  | **Unadjusted model** | | **Adjusted model** | |
| --- | --- | --- | --- | --- |
|  | **OR** | **95% CI** | **aOR** | **95% CI** |
| **Minority ethnicity status** |  |  |  |  |
| White British | ref | - | ref | - |
| Ethnic minority | 1.47 | 0.88, 2.41 | 1.27 | 0.74, 2.14 |
| **Age group** |  |  |  |  |
| 18-44 |  |  | ref | - |
| 45-64 |  |  | 0.54 | 0.33, 0.91 |
| 65+ |  |  | 0.59 | 0.35, 1.01 |
| **Sex-at-birth** |  |  |  |  |
| Male |  |  | ref | - |
| Female |  |  | 0.95 | 0.65, 1.39 |

**Table 20 Unadjusted odds ratio (OR) and adjusted odds ratio (aOR) for the association between ethnic minority status and experiencing limitations in taking care of others in the household.**

|  | **Unadjusted model** | | **Adjusted model** | |
| --- | --- | --- | --- | --- |
| Impact 4 | **OR** | **95% CI** | **aOR** | **95% CI** |
| **Minority ethnicity status** |  |  |  |  |
| White British | ref | - | ref | - |
| Ethnic minority | 0.75 | 0.41, 1.31 | 0.66 | 0.35, 1.17 |
| **Age group** |  |  |  |  |
| 18-44 |  |  | ref | - |
| 45-64 |  |  | 0.63 | 0.37, 1.06 |
| 65+ |  |  | 0.61 | 0.35, 1.06 |
| **Sex-at-birth** |  |  |  |  |
| Male |  |  | ref | - |
| Female |  |  | 1.17 | 0.8, 1.72 |

**Table 21 Unadjusted odds ratio (OR) and adjusted odds ratio (aOR) for the association between ethnic minority status and experiencing limitations in doing necessary activities in the house.**

|  | **Unadjusted model** | | **Adjusted model** | |
| --- | --- | --- | --- | --- |
|  | **OR** | **95% CI** | **aOR** | **95% CI** |
| **Minority ethnicity status** |  |  |  |  |
| White British | ref | - | ref | - |
| Ethnic minority | 0.84 | 0.53, 1.36 | 0.80 | 0.49, 1.31 |
| **Age group** |  |  |  |  |
| 18-44 |  |  | ref | - |
| 45-64 |  |  | 0.87 | 0.53, 1.41 |
| 65+ |  |  | 0.94 | 0.56, 1.55 |
| **Sex-at-birth** |  |  |  |  |
| Male |  |  | ref | - |
| Female |  |  | 1.36 | 0.98, 1.89 |

**Table 22 Unadjusted odds ratio (OR) and adjusted odds ratio (aOR) for the association between ethnic minority status and experiencing limitations in doing enjoyable activities.**

|  | **Unadjusted model** | | **Adjusted model** | |
| --- | --- | --- | --- | --- |
|  | **OR** | **95% CI** | **aOR** | **95% CI** |
| **Minority ethnicity status** |  |  |  |  |
| White British | ref | - | ref | - |
| Ethnic minority | 0.97 | 0.58, 1.66 | 0.92 | 0.54, 1.60 |
| **Age group** |  |  |  |  |
| 18-44 |  |  | ref | - |
| 45-64 |  |  | 0.80 | 0.45, 1.36 |
| 65+ |  |  | 0.90 | 0.50, 1.56 |
| **Sex-at-birth** |  |  |  |  |
| Male |  |  | ref | - |
| Female |  |  | 1.21 | 0.85, 1.72 |

Sensitivity analyses – Complete case analysis for IMD main analysis

**Table 23 Sensitivity Analyses - Unadjusted odds ratio (OR) and adjusted odds ratio (aOR) for the association between IMD quintile and experiencing limitations in attending or participating in work or education.**

|  | **Unadjusted model** | | **Adjusted model** | |
| --- | --- | --- | --- | --- |
|  | **OR** | **95% CI** | **aOR** | **95% CI** |
| **IMD Quintile** |  |  |  |  |
| 1 (most deprived) | 1.85 | 0.85, 4.21 | 1.68 | 0.76, 3.86 |
| 2 | 1.81 | 0.98, 3.38 | 1.66 | 0.89, 3.14 |
| 3 | 1.37 | 0.76, 2.50 | 1.30 | 0.71, 2.40 |
| 4 | 1.19 | 0.67, 2.14 | 1.25 | 0.68, 2.28 |
| 5 (least deprived) | ref | - | ref | - |
| **Age group** |  |  |  |  |
| 18-44 |  |  | ref | - |
| 45-64 |  |  | 0.94 | 0.52, 1.66 |
| 65+ |  |  | 0.47 | 0.24, 0.90 |
| **Sex-at-birth** |  |  |  |  |
| Male |  |  | ref | - |
| Female |  |  | 0.98 | 0.62, 1.54 |
| **Minority ethnicity status** |  |  |  |  |
| White British |  |  | ref | - |
| Ethnic minority |  |  | 0.80 | 0.35, 1.81 |
| **Migration status** |  |  |  |  |
| UK Born |  |  | ref | - |
| Not UK Born |  |  | 1.59 | 0.66, 3.96 |

**Table 24 Sensitivity Analyses - Unadjusted odds ratio (OR) and adjusted odds ratio (aOR) for the association between IMD quintile and experiencing limitations in concentrating.**

|  | **Unadjusted model** | | **Adjusted model** | |
| --- | --- | --- | --- | --- |
|  | **OR** | **95% CI** | **aOR** | **95% CI** |
| **IMD Quintile** |  |  |  |  |
| 1 (most deprived) | 3.22 | 1.52, 7.50 | 2.89 | 1.35, 6.79 |
| 2 | 1.75 | 1.03, 3.01 | 1.62 | 0.95, 2.82 |
| 3 | 1.41 | 0.86, 2.33 | 1.31 | 0.79, 2.19 |
| 4 | 1.05 | 0.65, 1.70 | 1.03 | 0.63, 1.67 |
| 5 (least deprived) | ref | - | ref | - |
| **Age group** |  |  |  |  |
| 18-44 |  |  | ref | - |
| 45-64 |  |  | 1.13 | 0.62, 2.03 |
| 65+ |  |  | 0.77 | 0.41, 1.39 |
| **Sex-at-birth** |  |  |  |  |
| Male |  |  | ref | - |
| Female |  |  | 1.25 | 0.85, 1.85 |
| **Minority ethnicity status** |  |  |  |  |
| White British |  |  | ref | - |
| Ethnic minority |  |  | 2.25 | 0.95, 5.79 |
| **Migration status** |  |  |  |  |
| UK Born |  |  | ref | - |
| Not UK Born |  |  | 0.66 | 0.27, 1.60 |

**Table 25 Sensitivity Analyses - Unadjusted odds ratio (OR) and adjusted odds ratio (aOR) for the association between IMD quintile and experiencing limitations in self-care.**

|  | **Unadjusted model** | | **Adjusted model** | |
| --- | --- | --- | --- | --- |
|  | **OR** | **95% CI** | **aOR** | **95% CI** |
| **IMD Quintile** |  |  |  |  |
| 1 (most deprived) | 1.97 | 0.88, 4.27 | 1.74 | 0.77, 3.85 |
| 2 | 2.11 | 1.14, 3.95 | 2.04 | 1.09, 3.85 |
| 3 | 2.05 | 1.14, 3.74 | 2.11 | 1.16, 3.89 |
| 4 | 2.09 | 1.17, 3.79 | 2.32 | 1.28, 4.27 |
| 5 (least deprived) | ref | - | ref | - |
| **Age group** |  |  |  |  |
| 18-44 |  |  | ref | - |
| 45-64 |  |  | 0.51 | 0.29, 0.91 |
| 65+ |  |  | 0.52 | 0.29, 0.97 |
| **Sex-at-birth** |  |  |  |  |
| Male |  |  | ref | - |
| Female |  |  | 0.82 | 0.53, 1.27 |
| **Minority ethnicity status** |  |  |  |  |
| White British |  |  | ref | - |
| Ethnic minority |  |  | 1.32 | 0.57, 2.96 |
| **Migration status** |  |  |  |  |
| UK Born |  |  | ref | - |
| Not UK Born |  |  | 1.16 | 0.48, 2.73 |

**Table 26 Sensitivity Analyses - Unadjusted odds ratio (OR) and adjusted odds ratio (aOR) for the association between IMD quintile and experiencing limitations in taking care of others in the household.**

|  | **Unadjusted model** | | **Adjusted model** | |
| --- | --- | --- | --- | --- |
|  | **OR** | **95% CI** | **aOR** | **95% CI** |
| **IMD Quintile** |  |  |  |  |
| 1 (most deprived) | 1.48 | 0.70, 3.08 | 1.41 | 0.66, 2.95 |
| 2 | 1.09 | 0.60, 1.98 | 1.05 | 0.57, 1.93 |
| 3 | 1.06 | 0.60, 1.87 | 1.05 | 0.59, 1.87 |
| 4 | 1.33 | 0.77, 2.31 | 1.27 | 0.73, 2.22 |
| 5 (least deprived) | ref | - | ref | - |
| **Age group** |  |  |  |  |
| 18-44 |  |  | ref | - |
| 45-64 |  |  | 0.6 | 0.33, 1.08 |
| 65+ |  |  | 0.62 | 0.33, 1.15 |
| **Sex-at-birth** |  |  |  |  |
| Male |  |  | ref | - |
| Female |  |  | 1.24 | 0.80, 1.94 |
| **Minority ethnicity status** |  |  |  |  |
| White British |  |  | ref | - |
| **Ethnic minority** |  |  | 0.95 | 0.38, 2.30 |
| Migration status |  |  |  |  |
| UK Born |  |  | ref | - |
| Not UK Born |  |  | 0.68 | 0.25, 1.76 |

**Table 27 Sensitivity Analyses - Unadjusted odds ratio (OR) and adjusted odds ratio (aOR) for the association between IMD quintile and experiencing limitations in doing necessary activities outside the house.**

|  | **Unadjusted model** | | **Adjusted model** | |
| --- | --- | --- | --- | --- |
|  | **OR** | **95% CI** | **aOR** | **95% CI** |
| **IMD Quintile** |  |  |  |  |
| 1 (most deprived) | 1.77 | 0.91, 3.55 | 1.77 | 0.91, 3.58 |
| 2 | 1.26 | 0.75, 2.12 | 1.27 | 0.76, 2.15 |
| 3 | 1.16 | 0.71, 1.88 | 1.14 | 0.70, 1.86 |
| 4 | 1.41 | 0.88, 2.29 | 1.35 | 0.83, 2.21 |
| 5 (least deprived) | ref | - | ref | - |
| **Age group** |  |  |  |  |
| 18-44 |  |  | ref | - |
| 45-64 |  |  | 0.86 | 0.49, 1.49 |
| 65+ |  |  | 0.98 | 0.54, 1.75 |
| **Sex-at-birth** |  |  |  |  |
| Male |  |  | ref | - |
| Female |  |  | 1.46 | 1.00, 2.12 |
| **Minority ethnicity status** |  |  |  |  |
| White British |  |  | ref | - |
| Ethnic minority |  |  | 0.70 | 0.32, 1.52 |
| **Migration status** |  |  |  |  |
| UK Born |  |  | ref | - |
| Not UK Born |  |  | 1.32 | 0.59, 3.05 |

**Table 28 Sensitivity Analyses - Unadjusted odds ratio (OR) and adjusted odds ratio (aOR) for the association between IMD quintile and experiencing limitations in doing enjoyable activities.**

|  | **Unadjusted model** | | **Adjusted model** | |
| --- | --- | --- | --- | --- |
|  | **OR** | **95% CI** | **aOR** | **95% CI** |
| **IMD Quintile** |  |  |  |  |
| 1 (most deprived) | 1.87 | 0.89, 4.24 | 1.90 | 0.90, 4.34 |
| 2 | 1.17 | 0.68, 2.03 | 1.19 | 0.69, 2.08 |
| 3 | 1.26 | 0.75, 2.14 | 1.28 | 0.76, 2.18 |
| 4 | 1.31 | 0.78, 2.22 | 1.33 | 0.79, 2.26 |
| 5 (least deprived) | ref | - | ref | - |
| **Age group** |  |  |  |  |
| 18-44 |  |  | ref | - |
| 45-64 |  |  | 0.85 | 0.45, 1.54 |
| 65+ |  |  | 1.05 | 0.54, 1.96 |
| **Sex-at-birth** |  |  |  |  |
| Male |  |  | ref | - |
| Female |  |  | 1.05 | 0.70, 1.57 |
| **Minority ethnicity status** |  |  |  |  |
| White British |  |  | ref | - |
| Ethnic minority |  |  | 0.84 | 0.37, 1.96 |
| **Migration status** |  |  |  |  |
| UK Born |  |  | ref | - |
| Not UK Born |  |  | 1.29 | 0.54, 3.20 |

**Sensitivity analyses**

**Table 29 Sensitivity Analyses - Unadjusted odds ratio (OR) and adjusted odds ratio (aOR) for the association between IMD quintile (without the health domain) and experiencing limitations in attending or participating in work or education.**

|  | **Unadjusted model** | | **Adjusted model** | |
| --- | --- | --- | --- | --- |
|  | **OR** | **95% CI** | **aOR** | **95% CI** |
| **IMD Quintile** |  |  |  |  |
| 1 (most deprived) | 2.31 | 1.15, 4.87 | 2.11 | 1.04, 4.48 |
| 2 | 2.39 | 1.37, 4.23 | 2.24 | 1.27, 4.00 |
| 3 | 1.26 | 0.76, 2.10 | 1.24 | 0.74, 2.09 |
| 4 | 1.16 | 0.69, 1.95 | 1.15 | 0.68, 1.94 |
| 5 (least deprived) | ref | - | ref | - |
| **Age group** |  |  |  |  |
| 18-44 |  |  | ref | - |
| 45-64 |  |  | 0.93 | 0.55, 1.56 |
| 65+ |  |  | 0.53 | 0.29, 0.94 |
| **Sex-at-birth** |  |  |  |  |
| Male |  |  | ref | - |
| Female |  |  | 0.98 | 0.65, 1.46 |
| **Minority ethnicity status** |  |  |  |  |
| White British |  |  | ref | - |
| Ethnic minority |  |  | 0.78 | 0.39, 1.55 |
| **Migration status** |  |  |  |  |
| UK Born |  |  | ref | - |
| Not UK Born |  |  | 1.51 | 0.67, 3.52 |
| Missing |  |  | 0.94 | 0.61, 1.46 |

**Table 30 Sensitivity Analyses - Unadjusted odds ratio (OR) and adjusted odds ratio (aOR) for the association between IMD quintile (without the health domain) and experiencing limitations in concentrating.**

|  | **Unadjusted model** | | **Adjusted model** | |
| --- | --- | --- | --- | --- |
|  | **OR** | **95% CI** | **aOR** | **95% CI** |
| **IMD Quintile** |  |  |  |  |
| 1 (most deprived) | 2.97 | 1.52, 6.20 | 2.80 | 1.43, 5.87 |
| 2 | 1.94 | 1.19, 3.20 | 1.87 | 1.14, 3.10 |
| 3 | 1.35 | 0.87, 2.09 | 1.34 | 0.86, 2.09 |
| 4 | 1.10 | 0.72, 1.69 | 1.13 | 0.73, 1.75 |
| 5 (least deprived) | ref | - | ref | - |
| **Age group** |  |  |  |  |
| 18-44 |  |  | ref | - |
| 45-64 |  |  | 0.96 | 0.55, 1.62 |
| 65+ |  |  | 0.68 | 0.38, 1.16 |
| **Sex-at-birth** |  |  |  |  |
| Male |  |  | ref | - |
| Female |  |  | 1.12 | 0.79, 1.58 |
| **Minority ethnicity status** |  |  |  |  |
| White British |  |  | ref | - |
| Ethnic minority |  |  | 1.42 | 0.72, 2.93 |
| **Migration status** |  |  |  |  |
| UK Born |  |  | ref | - |
| Not UK Born |  |  | 0.88 | 0.40, 1.98 |
| Missing |  |  | 1.24 | 0.84, 1.83 |

**Table 31 Sensitivity Analyses - Unadjusted odds ratio (OR) and adjusted odds ratio (aOR) for the association between IMD quintile (without the health domain) and experiencing limitations in self-care.**

|  | **Unadjusted model** | | **Adjusted model** | |
| --- | --- | --- | --- | --- |
|  | **OR** | **95% CI** | **aOR** | **95% CI** |
| **IMD Quintile** |  |  |  |  |
| 1 (most deprived) | 2.99 | 1.53, 5.87 | 2.79 | 1.41, 5.53 |
| 2 | 1.92 | 1.09, 3.40 | 1.85 | 1.04, 3.30 |
| 3 | 1.46 | 0.85, 2.55 | 1.48 | 0.85, 2.59 |
| 4 | 2.41 | 1.45, 4.10 | 2.49 | 1.49, 4.26 |
| 5 (least deprived) | ref | - | ref | - |
| **Age group** |  |  |  |  |
| 18-44 |  |  | ref | - |
| 45-64 |  |  | 0.59 | 0.35, 0.99 |
| 65+ |  |  | 0.66 | 0.39, 1.15 |
| **Sex-at-birth** |  |  |  |  |
| Male |  |  | ref | - |
| Female |  |  | 0.92 | 0.63, 1.36 |
| **Minority ethnicity status** |  |  |  |  |
| White British |  |  | ref | - |
| Ethnic minority |  |  | 1.2 | 0.59, 2.35 |
| **Migration status** |  |  |  |  |
| UK Born |  |  | ref | - |
| Not UK Born |  |  | 1.25 | 0.56, 2.72 |
| Missing |  |  | 1.05 | 0.68, 1.59 |

**Table 32 Sensitivity Analyses - Unadjusted odds ratio (OR) and adjusted odds ratio (aOR) for the association between IMD quintile (without the health domain) and experiencing limitations in taking care of others in the household.**

|  | **Unadjusted model** | | **Adjusted model** | |
| --- | --- | --- | --- | --- |
|  | **OR** | **95% CI** | **aOR** | **95% CI** |
| **IMD Quintile** |  |  |  |  |
| 1 (most deprived) | 1.81 | 0.93, 3.50 | 1.75 | 0.89, 3.42 |
| 2 | 1.56 | 0.91, 2.66 | 1.53 | 0.89, 2.63 |
| 3 | 0.90 | 0.53, 1.52 | 0.87 | 0.51, 1.47 |
| 4 | 1.60 | 0.99, 2.60 | 1.51 | 0.93, 2.47 |
| 5 (least deprived) | ref | - | ref | - |
| **Age group** |  |  |  |  |
| 18-44 |  |  | ref | - |
| 45-64 |  |  | 0.67 | 0.39, 1.14 |
| 65+ |  |  | 0.66 | 0.38, 1.17 |
| **Sex-at-birth** |  |  |  |  |
| Male |  |  | ref | - |
| Female |  |  | 1.17 | 0.79, 1.73 |
| **Minority ethnicity status** |  |  |  |  |
| White British |  |  | ref | - |
| Ethnic minority |  |  | 0.77 | 0.36, 1.58 |
| **Migration status** |  |  |  |  |
| UK Born |  |  | ref | - |
| Not UK Born |  |  | 0.76 | 0.30, 1.81 |
| Missing |  |  | 0.94 | 0.61, 1.44 |

**Table 33 Sensitivity Analyses - Unadjusted odds ratio (OR) and adjusted odds ratio (aOR) for the association between IMD quintile (without the health domain) and experiencing limitations in doing necessary activities outside the house.**

|  | **Unadjusted model** | | **Adjusted model** | |
| --- | --- | --- | --- | --- |
|  | **OR** | **95% CI** | **aOR** | **95% CI** |
| **IMD Quintile** |  |  |  |  |
| 1 (most deprived) | 2.03 | 1.11, 3.82 | 2.03 | 1.10, 3.85 |
| 2 | 1.80 | 1.12, 2.91 | 1.79 | 1.11, 2.91 |
| 3 | 1.07 | 0.70, 1.63 | 1.04 | 0.68, 1.59 |
| 4 | 1.45 | 0.95, 2.23 | 1.39 | 0.91, 2.14 |
| 5 (least deprived) | ref | - | ref | - |
| **Age group** |  |  |  |  |
| 18-44 |  |  | ref | - |
| 45-64 |  |  | 0.92 | 0.55, 1.50 |
| 65+ |  |  | 1.03 | 0.61, 1.72 |
| **Sex-at-birth** |  |  |  |  |
| Male |  |  | ref | - |
| Female |  |  | 1.36 | 0.98, 1.90 |
| **Minority ethnicity status** |  |  |  |  |
| White British |  |  | ref | - |
| Ethnic minority |  |  | 0.68 | 0.36, 1.28 |
| **Migration status** |  |  |  |  |
| UK Born |  |  | ref | - |
| Not UK Born |  |  | 1.27 | 0.61, 2.70 |
| Missing |  |  | 0.95 | 0.66, 1.38 |

**Table 34 Sensitivity Analyses - Unadjusted odds ratio (OR) and adjusted odds ratio (aOR) for the association between IMD quintile (without the health domain) and experiencing limitations in doing enjoyable activities.**

|  | **Unadjusted model** | | **Adjusted model** | |
| --- | --- | --- | --- | --- |
|  | **OR** | **95% CI** | **aOR** | **95% CI** |
| **IMD Quintile** |  |  |  |  |
| 1 (most deprived) | 2.05 | 1.04, 4.31 | 2.04 | 1.03, 4.30 |
| 2 | 1.41 | 0.86, 2.34 | 1.40 | 0.84, 2.33 |
| 3 | 1.16 | 0.73, 1.82 | 1.14 | 0.73, 1.81 |
| 4 | 1.49 | 0.94, 2.37 | 1.47 | 0.92, 2.34 |
| 5 (least deprived) | ref | - | ref | - |
| **Age group** |  |  |  |  |
| 18-44 |  |  | ref | - |
| 45-64 |  |  | 0.84 | 0.48, 1.45 |
| 65+ |  |  | 0.96 | 0.53, 1.69 |
| **Sex-at-birth** |  |  |  |  |
| Male |  |  | ref | - |
| Female |  |  | 1.20 | 0.84, 1.71 |
| **Minority ethnicity status** |  |  |  |  |
| White British |  |  | ref | - |
| Ethnic minority |  |  | 0.81 | 0.42, 1.63 |
| **Migration status** |  |  |  |  |
| UK Born |  |  | ref | - |
| Not UK Born |  |  | 1.26 | 0.57, 2.90 |
| Missing |  |  | 1.03 | 0.69, 1.55 |

**Table 35 Sensitivity Analyses - Unadjusted odds ratio (OR) and adjusted odds ratio (aOR) for the association between migration status (including the “Missing” category) and experiencing limitations in attending or participating in work or education.**

|  | **Unadjusted model** | | **Adjusted model** | |
| --- | --- | --- | --- | --- |
|  | **OR** | **95% CI** | **aOR** | **95% CI** |
| **Migration Status** |  |  |  |  |
| UK Born | ref | - | ref | - |
| Not UK Born | 1.55 | 0.82, 3.08 | 1.60 | 0.72, 3.72 |
| Missing | 0.86 | 0.57, 1.32 | 0.92 | 0.60, 1.41 |
| **Age group** |  |  |  |  |
| 18-44 |  |  | ref | - |
| 45-64 |  |  | 0.91 | 0.54, 1.51 |
| 65+ |  |  | 0.48 | 0.27, 0.85 |
| **Sex-at-birth** |  |  |  |  |
| Male |  |  | ref | - |
| Female |  |  | 1.01 | 0.68, 1.50 |
| **Minority Ethnicity Status** |  |  |  |  |
| White British |  |  | ref | - |
| Ethnic minority |  |  | 0.80 | 0.40, 1.57 |

**Table 36 Sensitivity Analyses - Unadjusted odds ratio (OR) and adjusted odds ratio (aOR) for the association between migration status (including the “Missing” category) and experiencing limitations in concentrating.**

|  | **Unadjusted model** | | **Adjusted model** | |
| --- | --- | --- | --- | --- |
|  | **OR** | **95% CI** | **aOR** | **95% CI** |
| **Migration Status** |  |  |  |  |
| UK Born | ref | - | ref | - |
| Not UK Born | 1.39 | 0.76, 2.66 | 0.95 | 0.44, 2.13 |
| Missing | 1.22 | 0.84, 1.79 | 1.20 | 0.82, 1.77 |
| **Age group** |  |  |  |  |
| 18-44 |  |  | ref | - |
| 45-64 |  |  | 0.91 | 0.53, 1.54 |
| 65+ |  |  | 0.60 | 0.35, 1.02 |
| **Sex-at-birth** |  |  |  |  |
| Male |  |  | ref | - |
| Female |  |  | 1.13 | 0.80, 1.59 |
| **Minority Ethnicity Status** |  |  |  |  |
| White British |  |  | ref | - |
| Ethnic minority |  |  | 1.45 | 0.74, 3.00 |

**Table 37 Sensitivity Analyses - Unadjusted odds ratio (OR) and adjusted odds ratio (aOR) for the association between migration status (including the “Missing” category) and experiencing limitations in self-care.**

|  | **Unadjusted model** | | **Adjusted model** | |
| --- | --- | --- | --- | --- |
|  | **OR** | **95% CI** | **aOR** | **95% CI** |
| **Migration Status** |  |  |  |  |
| UK Born | ref | - | ref | - |
| Not UK Born | 1.52 | 0.83, 2.71 | 1.27 | 0.58, 2.73 |
| Missing | 0.99 | 0.65, 1.49 | 0.99 | 0.65, 1.49 |
| **Age group** |  |  |  |  |
| 18-44 |  |  | ref | - |
| 45-64 |  |  | 0.54 | 0.33, 0.91 |
| 65+ |  |  | 0.59 | 0.35, 1.01 |
| **Sex-at-birth** |  |  |  |  |
| Male |  |  | ref | - |
| Female |  |  | 0.95 | 0.66, 1.39 |
| **Minority Ethnicity Status** |  |  |  |  |
| White British |  |  | ref | - |
| Ethnic minority |  |  | 1.12 | 0.56, 2.16 |

**Table 38 Sensitivity Analyses - Unadjusted odds ratio (OR) and adjusted odds ratio (aOR) for the association between migration status (including the “Missing” category) and experiencing limitations in taking care of others in the household.**

|  | **Unadjusted model** | | **Adjusted model** | |
| --- | --- | --- | --- | --- |
|  | **OR** | **95% CI** | **aOR** | **95% CI** |
| **Migration Status** |  |  |  |  |
| UK Born | ref | - | ref | - |
| Not UK Born | 0.71 | 0.34, 1.38 | 0.79 | 0.32, 1.87 |
| Missing | 0.90 | 0.59, 1.36 | 0.92 | 0.60, 1.39 |
| **Age group** |  |  |  |  |
| 18-44 |  |  | ref | - |
| 45-64 |  |  | 0.63 | 0.37, 1.06 |
| 65+ |  |  | 0.61 | 0.35, 1.05 |
| **Sex-at-birth** |  |  |  |  |
| Male |  |  | ref | - |
| Female |  |  | 1.17 | 0.80, 1.72 |
| **Minority Ethnicity Status** |  |  |  |  |
| White British |  |  | ref | - |
| Ethnic minority |  |  | 0.74 | 0.35, 1.51 |

**Table 39 Sensitivity Analyses - Unadjusted odds ratio (OR) and adjusted odds ratio (aOR) for the association between migration status (including the “Missing” category) and experiencing limitations in doing necessary activities outside the house.**

|  | **Unadjusted model** | | **Adjusted model** | |
| --- | --- | --- | --- | --- |
|  | **OR** | **95% CI** | **aOR** | **95% CI** |
| **Migration Status** |  |  |  |  |
| UK Born | ref | - | ref | - |
| Not UK Born | 1.03 | 0.59, 1.85 | 1.32 | 0.64, 2.81 |
| Missing | 0.90 | 0.63, 1.29 | 0.91 | 0.63, 1.31 |
| **Age group** |  |  |  |  |
| 18-44 |  |  | ref | - |
| 45-64 |  |  | 0.86 | 0.53, 1.41 |
| 65+ |  |  | 0.94 | 0.56, 1.56 |
| **Sex-at-birth** |  |  |  |  |
| Male |  |  | ref | - |
| Female |  |  | 1.37 | 0.99, 1.90 |
| **Minority Ethnicity Status** |  |  |  |  |
| White British |  |  | ref | - |
| Ethnic minority |  |  | 0.69 | 0.37, 1.29 |

**Table 40 Sensitivity Analyses - Unadjusted odds ratio (OR) and adjusted odds ratio (aOR) for the association between migration status (including the “Missing” category) and experiencing limitations in doing enjoyable activities.**

|  | **Unadjusted model** | | **Adjusted model** | |
| --- | --- | --- | --- | --- |
|  | **OR** | **95% CI** | **aOR** | **95% CI** |
| **Migration Status** |  |  |  |  |
| UK Born | ref | - | ref | - |
| Not UK Born | 1.13 | 0.61, 2.21 | 1.29 | 0.59, 2.95 |
| Missing | 0.99 | 0.67, 1.48 | 0.99 | 0.67, 1.49 |
| **Age group** |  |  |  |  |
| 18-44 |  |  | ref | - |
| 45-64 |  |  | 0.80 | 0.45, 1.36 |
| 65+ |  |  | 0.90 | 0.50, 1.57 |
| **Sex-at-birth** |  |  |  |  |
| Male |  |  | ref | - |
| Female |  |  | 1.21 | 0.85, 1.72 |
| **Minority Ethnicity Status** |  |  |  |  |
| White British |  |  | ref | - |
| Ethnic minority |  |  | 0.80 | 0.41, 1.60 |
